# How intense is effective? Exploring aerobic exercise intensity for knee osteoarthritis through a Bayesian network meta-analysis

**DOI:** 10.1101/2025.09.08.25335181

**Authors:** Luis Cabrera-Durán, Javier Palomares-Fernández, Ignacio Canitrot-González, Paride Crisafulli, Carlos Donato Cabrera-López, José Fierro-Marrero

## Abstract

**Introduction:** Knee osteoarthritis (KOA) is a degenerative disease of the articular cartilage, characterized by pain, stiffness, and functional limitation. Exercise is one of the main treatment options. Aerobic exercise has shown favorable effects in these patients.

**Aims:** Determine which intensity of aerobic exercise (AE) is most effective for these patients.

**Methods:** Searches were conducted in 7 databases, complemented by citation and manual searches. Eligible studies were randomized controlled trials with an AE-only group. Outcome measures were knee pain intensity, walking performance, sit-to-stand performance, combined sit-to-stand and walking performance, perceived knee stiffness and disability. Methodological quality and risk of bias were assessed. Two Bayesian random-effects network meta-analysis models were employed for comparing different AE intensities, reporting standardized mean differences (Hedges’ g) with 95% credible intervals. The certainty of evidence was evaluated using the adapted GRADE framework for network meta-analysis exploring risk of bias, indirectness, heterogeneity, publication bias, intransitivity, incoherence and imprecision of the estimate.

**Results:** Fifteen studies were finally included. They had an average of 5.93 points in the PEDro scale and a risk of bias including “some concerns” and “high risk”. Two meta-analyses (pain intensity and walking performance) were conducted. Comparisons between AE intensities presented non-significant and imprecise results, classified with a very low certainty of evidence.

**Conclusions:** Very low certainty of evidence prevents clear recommendations on the optimal dose of AE. Current findings are limited by high risk of bias, imprecise credible intervales, and reliance on indirect comparisons. Clinicians should strictly individualize intensity based on patient characteristics, preferences, and clinical response, and combine AE with other interventions (physiotherapy, self-management, education, medication control). Medium-to-long term AE programs are advisable given their additional benefits on frequent comorbidities. More high-quality trials directly comparing AE intensities are needed.

## 1. Introduction

According to the Osteoarthritis Research Society International, osteoarthritis (OA) is a disease of the articular cartilage, in which it undergoes progressive degradation. OA first manifests as a molecular disorder (abnormal joint tissue metabolism) followed by observable anatomical and/or physiological disorders, such as, articular cartilage degradation, bone remodeling, osteophyte formation, joint inflammation, and loss of normal joint function (Kraus et al., 2015). The intensity of OA symptoms varies, but they typically worsen over time. The most common ones tend to be pain, stiffness, and swelling of the knee. The severity of the disease can be classified based on radiological findings using the Kellgren-Lawrence system (KELLGREN & LAWRENCE, 1957).

KOA presents a global prevalence of 22.9% of individuals over 40 years, being examined through both radiological and symptomatic-radiological diagnosis (Cui et al., 2020). The global incidence rate is 203 per 10,000 person-years among individuals aged 20 and above, with higher rates observed for radiological diagnosis compared to symptomatic diagnosis. Both prevalence and incidence rates increase with age, and notably, both are higher in women (Cui et al., 2020). A study conducted in 2019 estimated an average annual cost of KOA at 2,295 € per patient, with expenses increasing along with the severity of joint degeneration (Salmon et al., 2019). In the United States, annual costs can reach up to 10,000 € per patient, a difference potentially attributed to the higher prevalence of obesity in this country (Salmon et al., 2016).

Current guidelines recommend a multimodal approach to manage OA, incorporating interventions such as exercise, education, and weight management (Gibbs et al., 2023). Within this framework, the literature supports the clinical benefits of exercise for OA management, emphasizing the prescription of therapeutic exercise (Uthman et al., 2014). Exercise is believed to produce these clinical benefits, by targeting different pathophysiological pathways involved in OA. Evidence from animal models suggests that exercise reduces the degradation of the articular cartilage, synovial membrane and subchondral bone, by enhancing autophagy, reducing apoptosis, increasing the production of extracellular matrix components, and lowering pro-inflammatory markers (Kong et al., 2022).

Exercise is a cost-effective intervention widely studied. However, there remains insufficient evidence to determine the optimal modality and dosage for exercise prescription in KOA. In the last decades, research has increasingly focused on identifying the most effective exercise modality, including interventions such as aerobic, mind-body, resistance, flexibility or any combination of these. While all of these modalities demonstrate efficacy compared to usual care among different clinical and performance outcomes, evidence suggests no relevant differences in efficacy among these modalities in knee and hip OA (Goh et al., 2019).

On the other hand, few studies have investigated the effects of exercise dosage parameters in this population. In this context, the FITT-VP model proposed by the American College of Sports Medicine (American College of Sports Medicine et al., 2022) provides a valuable framework for structuring exercise dosage. It is important to note that because exercise modalities differ significantly, dosage parameters should be examined individually for each modality.

For example, a study by Juhl et al. (2014), explored the effects of exercise modality and various dosage parameters in KOA. These parameters included exercise intensity, session duration, weekly frequency, number of weeks, and number of supervised sessions. Among these, the number of supervised sessions was positively associated with clinical improvements in some modalities, such as aerobic exercise. In contrast, no specific dosage parameters were found to significantly influence outcomes for resistance exercise. Similarly, findings from related meta-analysis suggest that exercise intensity, whether in aerobic or resistance modalities, do not lead to clinically significant differences (Regnaux et al., 2015).

Aerobic exercise is defined as an exercise intervention that uses large muscle groups, with increased breathing and continuously maintaining a heart rate at a determinate intensity (American College of Sports Medicine et al., 2022). The effects of aerobic exercise on cardiovascular health and systemic inflammation are well-established (Zheng et al., 2019), and produces clinical benefits in patients with KOA (Goh et al., 2019). However, the best recommendations for prescribing aerobic exercise remain unclear. To our knowledge, no meta-analysis has been conducted exploring the clinical effectiveness of aerobic exercise (isolated) through its different intensities of prescription, in patients with KOA. Therefore, based on the present state of the art, we consider conducting such study to provide valuable insights on the prescription of aerobic exercise in this population.

## 2. Methods

This systematic review and network-meta-analysis (NMA) followed the Preferred Reporting Items for Overviews of Systematic Reviews and Meta-analysis Network Meta-analysis Extension Statement (Hutton et al., 2015). The protocol was also registered in PROSPERO (CRD42024519632).

### 2.1. Selection criteria

Selection criteria were based on the PICOS strategy.

#### 2.1.1. Population

Patients with KOA were eligible regardless of the diagnostic criteria applied. No exclusions were made based on demographic variables (age, sex, etc.). Studies including patients with KOA and other concomitant musculoskeletal pathologies, such as hip OA, were excluded.

#### 2.1.2. Intervention

Eligible interventions must consist solely of isolated aerobic exercise. Studies which added any type of intervention, lifestyle recommendation or educational issues were excluded. Studies should include internal load parameters of aerobic exercise intensity, such as heart rate (HR), oxygen consumption measures, or subjective internal parameters like perceived exertion or talk test.

#### 2.1.3. Comparison

Any type of comparator was eligible for inclusion. Comparison groups would serve as connector nodes in the NMA for estimating indirect evidence between aerobic exercise interventions.

#### 2.1.4. Outcome measures and time-point analysis

The following outcome measures were considered for inclusion:

1. Knee pain intensity: Any instrument exploring this outcome measure was considered for inclusion.
2. Performance in walking tasks: Gait tasks based on a limited distance or time were eligible for inclusion.
3. Performance in sit-to-stand tasks: Sit-to-stand tasks based on a limited repetitions or time were eligible for inclusion.
4. Performance in combined sit-to-stand and walking tasks: Tests like the Timed-up-and-go (TUG) or similar were eligible.
5. Perceived knee stiffness: Any instrument exploring this outcome measure was considered for inclusion.
6. Disability related with KOA: The only eligible tool for this outcome measure was the total score in Western Ontario McMaster Universities Osteoarthritis Index (WOMAC).

Only immediate evaluations after treatment were eligible. Other time-points, such as during-intervention and follow-up evaluations were excluded.

#### 2.1.5. Data availability

Eligible studies should present sample size, central tendency parameters (mean or median) and dispersion parameters (quartiles, interquartile range, standard deviation, standard error of the mean, confidence intervals) for the outcomes and time-points of interest.

#### 2.1.6. Study design

Only randomized controlled trials were eligible study designs.

### 2.2. Search strategy

Systematic searches were conducted using PubMed, EBSCO, Web of Science, SciELO, ScienceDirect, Scopus, and Google Scholar to find eligible studies across multiple databases. Search engines, search equations, databases, number of registries retrieved, and search dates are reported in Supplementary Material. Additionally, non-systematic manual and citation searches were performed.

### 2.3. Selection process

The selection process was conducted by two researchers (LCD and ICG) using Rayyan.ai (Ouzzani et al., 2016). Duplicate records were identified automatically and manually deleted collaboratively. After duplicate removal, both reviewers independently and blindly selected studies for inclusion.

The selection process had two stages. In the first stage, reviewers evaluated Title-Abstract-Keywords-Design to decide whether to exclude the record or advance it to Full-Text analysis. Exclusions were only conducted if clear information supported this decision. A pilot test, was conducted with the first 60 registries to refine the decision-making process. Initially, the first 30 records were analyzed, and reviewers’ decisions were compared to those of the research group. Following group discussion, reviewers reanalysed the initial 30 records and prceeded with records 31-60, again comparing their decisions with the group’s conclusions.

After completing the Title-Abstract-Keywords-Design screening, registries agreed upon for exclusion were removed, while those agreed upon for Full-Text analysis were advanced to the next stage. Disagreements between reviewers were resolved between reviewers, with assistance from a third reviewer.

In the second stage, records selected for Full-Text analysis were reassessed. Records agreed upon for inclusion were included in the study, while those agreed upon for exclusion were removed. Discrepancies were resolved between reviewers, with assistance from a third reviewer.

The selection process for manual and citation searches was conducted collaboratively by the reviewers.

### 2.4. Data extraction

#### 2.4.1. Summary information

The summary information would include authors, study design, KOA diagnostic criteria and the presence of pain. Additionally, the BMI, diabetes, blood pressure values would be provided. Experimental (aerobic) and control groups (other interventions), allocated sample size, age (mean and SD), and sex (female sample size) within groups would be provided.

Outcome measures eligible for extraction would include: 1) knee pain intensity; 2) performance in walking tasks; 3) performance in sit-to-stand tasks; 4) performance in combined tasks of sit-to-stand and walking tasks; 5) perceived knee stiffness, and 6) disability related to KOA. Their assessment tools would be reported. Immediately-after the intervention results would be provided for the comparisons between experimental groups, and experimental with control groups. This would be presented narratively and summarized indicating the direction of the effect (>, <, or ≈ between groups).

#### 2.4.2. Exercise prescription

Aerobic exercise prescription parameters were extracted. These include the exercise activity (walking, cycling, etc.), and density features (intervallic or continuous modalities), In addition, volume parameters would be reported, including:

- Total session duration (including exercise and rest periods).
- Aerobic exercise stimuli duration (specific time exercising).
- Weekly frequency.
- Number of weeks.

Additionally, the intensity of aerobic exercise, monitorization instruments, and the intensity progression parameters would be reported. Exercise intensity parameters would be categorized into “near-maximal”, “vigorous”, “moderate”, “light”, “very light”, or any of their combinations (such as very light-to-light, etc.) following the criteria by the American College of Sports Medicine (American College of Sports Medicine et al., 2022) for percentage of maximum heart rate (%HRmax), percentage of maximum heart rate reserve (%HRR), percentage of maximal oxygen consumption (%VO_2_max), percentage of oxygen consumption reserve (%VO_2_reserve), and Borg 6-20 scale, see Supplementary Material. Estimations for other instruments such as the Borg CR-10 would be conducted based on the conversions proposed by (Borg, 1998). Talk test zones would be identified (Foster et al., 2018), to estimate HR, or oxygen consumption measures, and consequently the intensity (Dubiel, 2014).

#### 2.4.3. Data extraction procedure

Three researchers (ICG, JPF and JFM) independently and blindly extracted the following information: 1) type of intervention in the experimental and control groups; 2) aerobic exercise intensity prescription data; 3) categorization of exercise intensity into “near-maximal”, “vigorous”, “moderate”, “light”, “very light”, or their combinations; 4) outcome measures of interest, and the instruments/tests employed; 5) sample size, central tendency and dispersion results of the outcomes of interest immediately after the intervention.

The agreed information was considered valid, included in the summary table, and used for meta-analyses. Disagreements would be resolved between reviewers. The remaining information was extracted by LCD.

### 2.5. Methodological quality and risk of bias

Two reviewers (LCD and JPF) would blindly and independently assess the methodological quality using the PEDro scale and the risk of bias using the Risk of Bias Tool 2.0 for the randomized controlled trials included in the review (Morton, 2009; Sterne et al., 2019).

The PEDro scale consists of 11 items: (1) eligibility criteria, (2) randomization, (3) concealment, (4) baseline similarity between groups, (5) blinding of participants, (6) blinding of therapists, (7) blinding of assessors, (8) <15% dropouts, (9) “intention-to-treat” analysis of the dropouts, (10) between-groups analysis available, (11) Point and variability measures reported. Each item receives one point if the answer is “yes”; a “no” answer gets 0 points. Items 2 to 11 are used to calculate the total score, which ranges from 0 to 10 points.

The ROB 2.0 assesses risk of bias through five domains: (1) randomization process and (2) deviations from the intended intervention were assessed once for each study. The remaining domains were evaluated for every outcome measure of interest. These include (3) missing outcome data, (4) outcome measurement and (5) selection of the reported results. Each question was evaluated using questions with five possible answers: “yes”, “probably yes”, “probably no”, “no” or “no information”. Based on these answers and the algorithms proposed in the scale, a risk of bias is assigned to each domain with three possible outcomes: “low risk”, “some concerns” or “high risk”. If all domains present “low risk”, the study is categorized as “low risk”. If at least one domain presents “some concerns” but not “high risk”, the study is categorized as having “some concerns”. If at least one domain presents a “high risk” of bias, this study is categorized as “high risk” of bias. Additionally, if 4 or more domains presented “some concerns”, the final risk of bias is considered “high risk”.

Inter-rater items’ level of agreement for each study and all studies combined was assessed using a Quadratic Weighted Cohen’s Kappa Coefficient (κ), with the following interpretation: “almost perfect” agreement: 0.81–1.00, “substantial” if 0.61–0.80, “moderate” if 0.41–0.6, “fair” if 0.21–0.4, “slight” if 0.00–0.20 and “poor”□when <□0.00 (Landis & Koch, 1977). Cohen’s Kappa was calculated using the “cohen.kappa” function from the “psych” package version 2.3.12 (William Revelle, 2024) in R Software version 4.3.1 (R Core Team, 2023). Agreed items would be maintained, while disagreements were resolved among group’s decision.

### 2.6. Network meta-analysis

A network meta-analysis (NMA) was conducted to compare the efficacy of aerobic exercise across different intensity levels, with “no treatment” serving as the primary control node. Experimental nodes represented specific aerobic exercise intensities classified as near-maximal, vigorous, moderate, light, very light, or any combination of these, following previously established methodology.

Standardized mean differences (SMDs) were calculated using Hedges’ g (Hedges, 1982) and interpreted as follows: very small (<0.20), small (0.20–0.49), medium (0.50– 0.79), and large (≥0.80) (Cohen, 1992). This calculation is specified in Supplementary Material.

Two Bayesian NMA models were implemented:

- Model 1: A Bayesian model including only the intervention effect (no covariates).
- Model 2: An adjusted Bayesian model including the intervention effect plus two study-level covariates: intervention duration (number of weeks) and weekly frequency.

The Bayesian hierarchical NMA was implemented using a custom Python script (see Supplementary Material), whose main class is based on the PyMC3 module (Salvatier et al., 2016). The primary analysis modelled different aerobic exercise intensities as nodes in the network. Additionally, based on available data from the original studies, a second NMA was conducted incorporating the covariates weekly frequency (w_f_), and number of weeks (w_n_). The relative efficacy, d_bi_, of treatment *i* was modeled as:

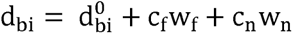

where transitivity holds for baseline estimates 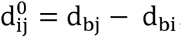, and coefficients c_f_ and c_w_ are inferred from the model. For non-aerobic treatments, both w_f_ and w_n_ were set to zero. Trial-specific efficacy Δ_t,ij_ was sampled from a normal distribution N(d_ij_, τ^2^), where τ\tauτ is the heterogeneity parameter. Patient-specific efficacy was then sampled from N(d_ij_, τ^2^) where 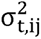 is the trial-specific variance extracted from the data. These two levels of randomness make the model a Random Effects model.

The prior distribution for each d_bi_ was a normal distribution centred at zero with variance of 100. For τ, we used a uniform distribution over [0, 5]. Both priors were chosen to be non-informative.

The Bayesian models would produced three main outputs:

1. Network graph indicating the direct connections established within each NMA.
2. Forest plot comparing exercise interventions against the no treatment control.
3. Pairwise comparison between interventions, reported as network (mixed), direct and indirect estimates. Inconsistency between direct and indirect evidence was tested with a two-tailed z-test.
4. Funnel plots to assess potential publication bias.

For both the forest plots and the pairwise comparisons, results were expressed as Hedges’ g, with the corresponding 95% credible intervals (CrI), p-values, and τ estimates. Direct estimates were obtained via a weighted average of all available direct comparisons, where the weights were the inverse of the corresponding standard errors.

To evaluate the potential for publication bias and inconsistency within the network, we generated comparison-adjusted funnel plots for each outcome. For each pairwise comparison between treatments within a study, we calculated the difference between the study’s observed effect size and the corresponding effect estimate from the NMA model. The standard error of this difference was plotted on the y-axis. In the absence of significant bias or inconsistency, the data points are expected to lie within the 95%CrI of the funnel, which is bounded by the lines x = ± 1.96 * SE. Points located outside this funnel indicate a greater-than-expected discrepancy between the direct and network estimates, which can be indicative of publication bias, heterogeneity, or other systematic sources of inconsistency. The data points are colored by study and shaped to denote the type of intervention comparison: triangles for comparisons between two AE interventions, circles for comparisons between two non-AE interventions, and squares for mixed comparisons between an AE and a non-AE intervention.

### 2.7. Synthesis of results

The synthesis of results will be presented with the adapted Cochrane GRADE (Grading of Recommendations Assessment, Development and Evaluation) of evidence for Network meta-analysis (Izcovich et al., 2023). This framework allows drawing conclusions on the certainty of the available evidence provided by the NMA, based on a stepwise evaluation of direct, indirect and mixed evidence.

For direct evidence, certainty is assessed across four GRADE domians: risk of bias, heterogeneity, indirectness and publication bias.

For indirect evidence, certainty ratings are derived from the direct comparisons forming the most dominant first or second order loop. The starting point is the lowest certainty rating of those contributing direct comparisons of the loop, testing for the same previous GRADE domains. In addition, indirect evidence is assessed for intransitivity.

For mixed (NMA) evidence, the starting point is the certainty rating of either the direct or indirect evidence, depending on which provides the most dominant contribution in the NMA. The mixed estimate is then further assessed for incoherence (tested through a z-test between direct and indirect estimates), and for imprecision (a mandatory stage, based on the width of the 95%CrI relative to decision-making thresholds).

Each domain is judged as “not serious”, “serious”, or “very serious”, leading to downgrading the certainty of evidence by 0, 1 or 2 levels respectively. Limitations across domains result in an overall rating of the certainty of evidence as high, moderate, low or very low.

## 3. Results

### 3.1. Selection process

A total of 6134 records were retrieved from the databases. After removing duplicates, 3216 records were assessed for eligibility by analyzing Title, Abstract, Keywords, and Study design. After resolving discrepancies, 207 records were analyzed in full text. A total of 15 studies (Arrieiro et al., 2019; Bavardi Moghadam & Shojaedin, 2017; Beckwée et al., 2015; Casilda-López et al., 2017; de Almeida et al., 2019, 2020; Ettinger et al., 1997; Keogh et al., 2018; Lim et al., 2010; Mangione et al., 1999; Messier et al., 1997; Øiestad et al., 2023; Salacinski et al., 2012; Samut et al., 2015; Watanabe & Someya, 2013) were finally included in the review after solving disagreements. See Figure 1 for further details of the selection process.

**Figure 1.**
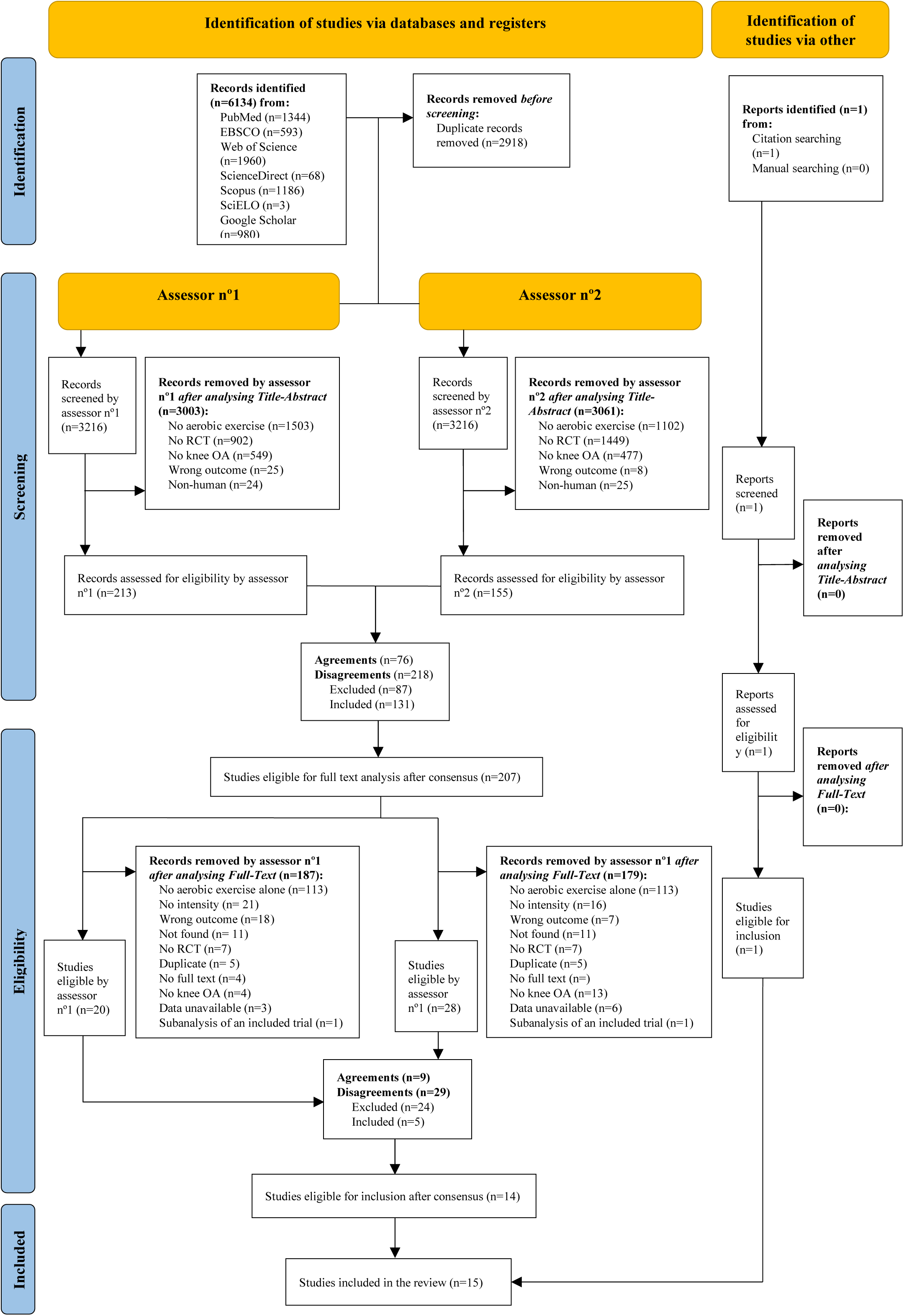
Flowchart of search and selection process

### 3.2. Summary information

Subjects across trials were recruited based on different diagnostic criteria, with 12 studies selecting patients with both clinical and radiological criteria (Arrieiro et al., 2019; Bavardi Moghadam & Shojaedin, 2017; Beckwée et al., 2015; de Almeida et al., 2019, 2020; Ettinger et al., 1997; Mangione et al., 1999; Messier et al., 1997; Øiestad et al., 2023; Salacinski et al., 2012; Samut et al., 2015; Watanabe & Someya, 2013), only clinical (Casilda-López et al., 2017), only radiological (Lim et al., 2010), or with an unstandardized procedure (diagnosed by a surgeon) (Keogh et al., 2018).

The presence of pain was explicitly confirmed in 9, which included patients with knee pain based on their selection criteria (Bavardi Moghadam & Shojaedin, 2017; Beckwée et al., 2015; de Almeida et al., 2019, 2020; Ettinger et al., 1997; Mangione et al., 1999; Messier et al., 1997; Øiestad et al., 2023; Salacinski et al., 2012). In 5 other studies, the presence of pain was likely but not clearly stated, as it was inferred from baseline pain assessments (Arrieiro et al., 2019; Casilda-López et al., 2017; Lim et al., 2010; Samut et al., 2015; Watanabe & Someya, 2013). In one study, the presence of knee pain was completely unclear (Keogh et al., 2018).

A total of 455 patients were assigned to aerobic exercise groups. Among them, 30 patients participated in a very light-to-light intensity aerobic exercise (Watanabe & Someya, 2013); 20 were assigned to light-to-moderate intensity exercise (Mangione et al., 1999); 22 were assigned to light-to-moderate-to-vigorous exercise (de Almeida et al., 2019, 2020); 31 participated in moderate intensity exercise (Keogh et al., 2018; Salacinski et al., 2012); 235 were assigned to a moderate-to-vigorous intensity exercise (Bavardi Moghadam & Shojaedin, 2017; Ettinger et al., 1997; Lim et al., 2010; Messier et al., 1997; Øiestad et al., 2023); and 117 engaged vigorous intensity exercise (Arrieiro et al., 2019; Beckwée et al., 2015; Casilda-López et al., 2017; Keogh et al., 2018; Mangione et al., 1999; Samut et al., 2015).

A total of 576 subjects were assigned to control groups. Among these, 259 subjects were allocated to resistance exercise programs (Beckwée et al., 2015; de Almeida et al., 2019, 2020; Ettinger et al., 1997; Messier et al., 1997; Øiestad et al., 2023; Samut et al., 2015); 171 to education-based interventions (de Almeida et al., 2019, 2020; Ettinger et al., 1997; Messier et al., 1997); 25 to a combined exercise program (Lim et al., 2010); 24 to resistance exercise combined with behavioural and lifestyle modification strategies (Lim et al., 2010); and 97 to no-treatment groups (Bavardi Moghadam & Shojaedin, 2017; Øiestad et al., 2023; Salacinski et al., 2012; Samut et al., 2015).

Aerobic exercise intensity was monitored using several procedures across studies. These included analysis of %HRmax (Arrieiro et al., 2019; Bavardi Moghadam & Shojaedin, 2017; de Almeida et al., 2019, 2020; Lim et al., 2010; Øiestad et al., 2023; Salacinski et al., 2012; Watanabe & Someya, 2013), %HRR (Ettinger et al., 1997; Mangione et al., 1999; Messier et al., 1997; Samut et al., 2015), the talk test (Keogh et al., 2018), the Borg Rating of Perceived Exertion (6-20 points) (Beckwée et al., 2015; de Almeida et al., 2019, 2020; Watanabe & Someya, 2013), or the Borg CR10 scale (Casilda-López et al., 2017).

The number of weeks for aerobic exercise protocols ranged from 6 to 79 weeks. The mean duration was 16.9 ± 21.5, with a median of 10 weeks (Q1= 8, Q3=13). The most frequent duration was 8 weeks. The weekly frequency of aerobic exercise sessions was 2.93 ± 0.53 with a median and mode of 3 sessions per week (Q1=3, Q3=3), ranging from 3 to 4 session per week.

Out of the 15 included articles, 12 measured pain intensity (Arrieiro et al., 2019; Beckwée et al., 2015; Casilda-López et al., 2017; de Almeida et al., 2019, 2020; Ettinger et al., 1997; Lim et al., 2010; Messier et al., 1997; Øiestad et al., 2023; Salacinski et al., 2012; Samut et al., 2015; Watanabe & Someya, 2013). The tools used to assess pain included the WOMAC pain subscale (Arrieiro et al., 2019; Casilda-López et al., 2017; de Almeida et al., 2019; Samut et al., 2015), the numerical pain rating scale (Ettinger et al., 1997; Øiestad et al., 2023), the brief pain intensity scale (Lim et al., 2010), the visual analogue scale (de Almeida et al., 2019; Salacinski et al., 2012; Samut et al., 2015; Watanabe & Someya, 2013) and a 1-6 point scale (Beckwée et al., 2015; Messier et al., 1997).

Walking performance was assessed in 10 studies (Arrieiro et al., 2019; Bavardi Moghadam & Shojaedin, 2017; Casilda-López et al., 2017; de Almeida et al., 2019; Ettinger et al., 1997; Keogh et al., 2018; Mangione et al., 1999; Salacinski et al., 2012; Samut et al., 2015; Watanabe & Someya, 2013) using a variety of tests: the 6-minute walk test (Arrieiro et al., 2019; Bavardi Moghadam & Shojaedin, 2017; Casilda-López et al., 2017; Ettinger et al., 1997; Mangione et al., 1999; Samut et al., 2015; Watanabe & Someya, 2013), the 40-m walk test (de Almeida et al., 2019), the 22.5-m walk test (Messier et al., 1997), the 10-m walk test (Watanabe & Someya, 2013), a 3.87-m walk at normal and maximal speed (Mangione et al., 1999), and a 3.66-m walk test at normal (Keogh et al., 2018; Salacinski et al., 2012) and maximal speed (Salacinski et al., 2012).

Sit-to-stand performance was evaluated in 4 studies (de Almeida et al., 2019; Keogh et al., 2018; Mangione et al., 1999; Samut et al., 2015), using either the 30-s sit-to-stand test (de Almeida et al., 2019; Keogh et al., 2018; Samut et al., 2015), or the 10-repetition chair-to-stand test (Mangione et al., 1999).

Three studies assessed tests which combined walking and sit-to-stand performance (Bavardi Moghadam & Shojaedin, 2017; Keogh et al., 2018; Watanabe & Someya, 2013) employing the timed-up-and-go test (Keogh et al., 2018; Watanabe & Someya, 2013), and a combination of a chair-to-stand with 15.2 m walk (Bavardi Moghadam & Shojaedin, 2017).

Perceived knee stiffness was analysed in 5 studies using the WOMAC stiffness subscale (Arrieiro et al., 2019; Casilda-López et al., 2017; de Almeida et al., 2019; Salacinski et al., 2012; Samut et al., 2015).

Disability associated with KOA was assessed in 5 studies using the total WOMAC score (Casilda-López et al., 2017; de Almeida et al., 2019; Keogh et al., 2018; Salacinski et al., 2012; Samut et al., 2015). See Table 1 for further details of the included studies.

### 3.3. Methodological quality and risk of bias

Of the 15 evaluated studies, the average score on the PEDro scale was 5.93 points (ranging from 4 to 8). Inter-rater agreement ranged from moderate (κ = 0.55) to perfect (κ = 1), see Figure 2.

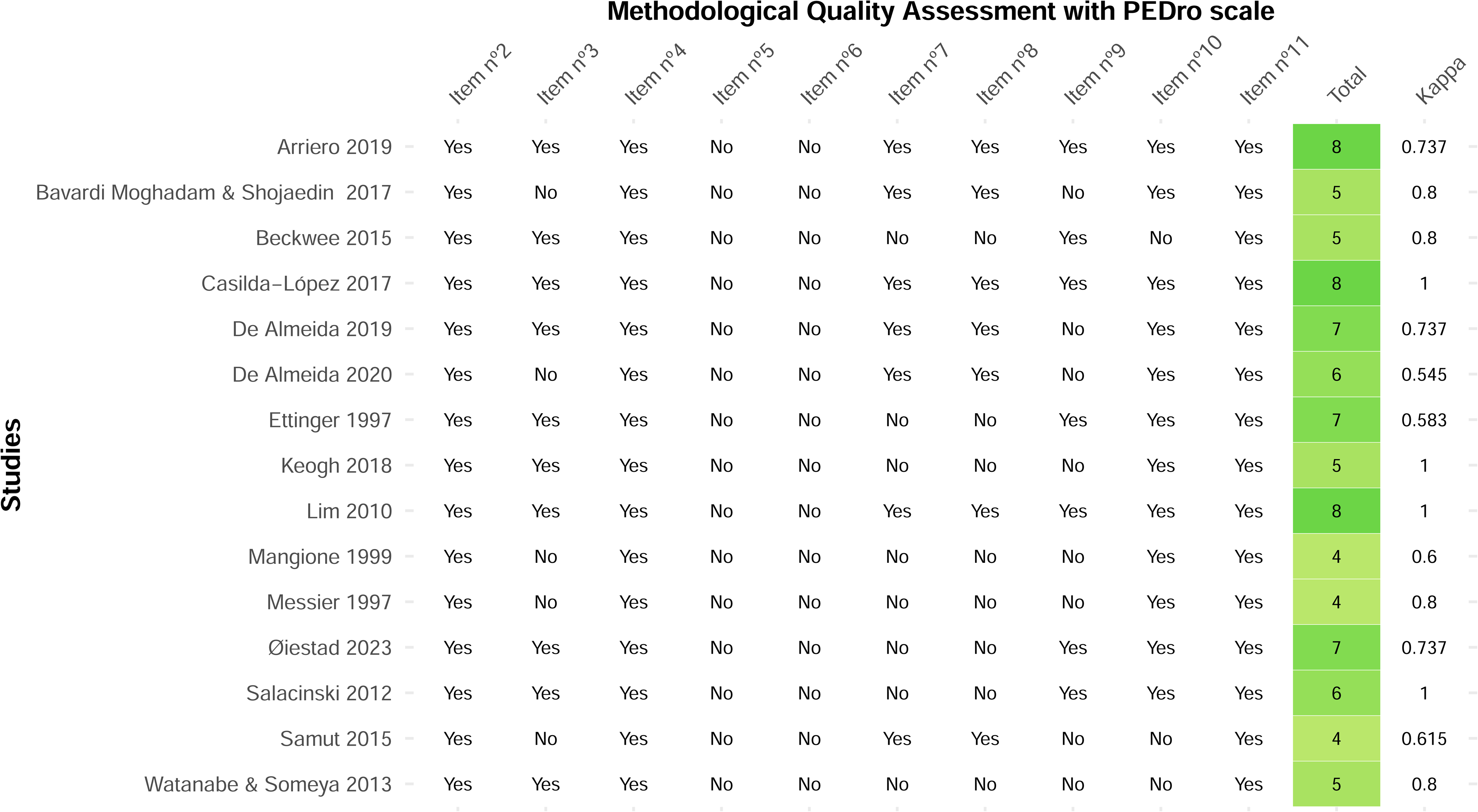

Regarding the ROB 2.0 scale, for the pain intensity outcome, 8 articles were rated as having “high risk” of bias (Arrieiro et al., 2019; Beckwée et al., 2015; Ettinger et al., 1997; Lim et al., 2010; Messier et al., 1997; Salacinski et al., 2012; Samut et al., 2015; Watanabe & Someya, 2013), and 3 presented “some concerns” (Casilda-López et al., 2017; de Almeida et al., 2019; Øiestad et al., 2023). Inter-rater agreement for this outcome ranged from slight (κ = 0) to perfect (κ = 1), see Figures 3.

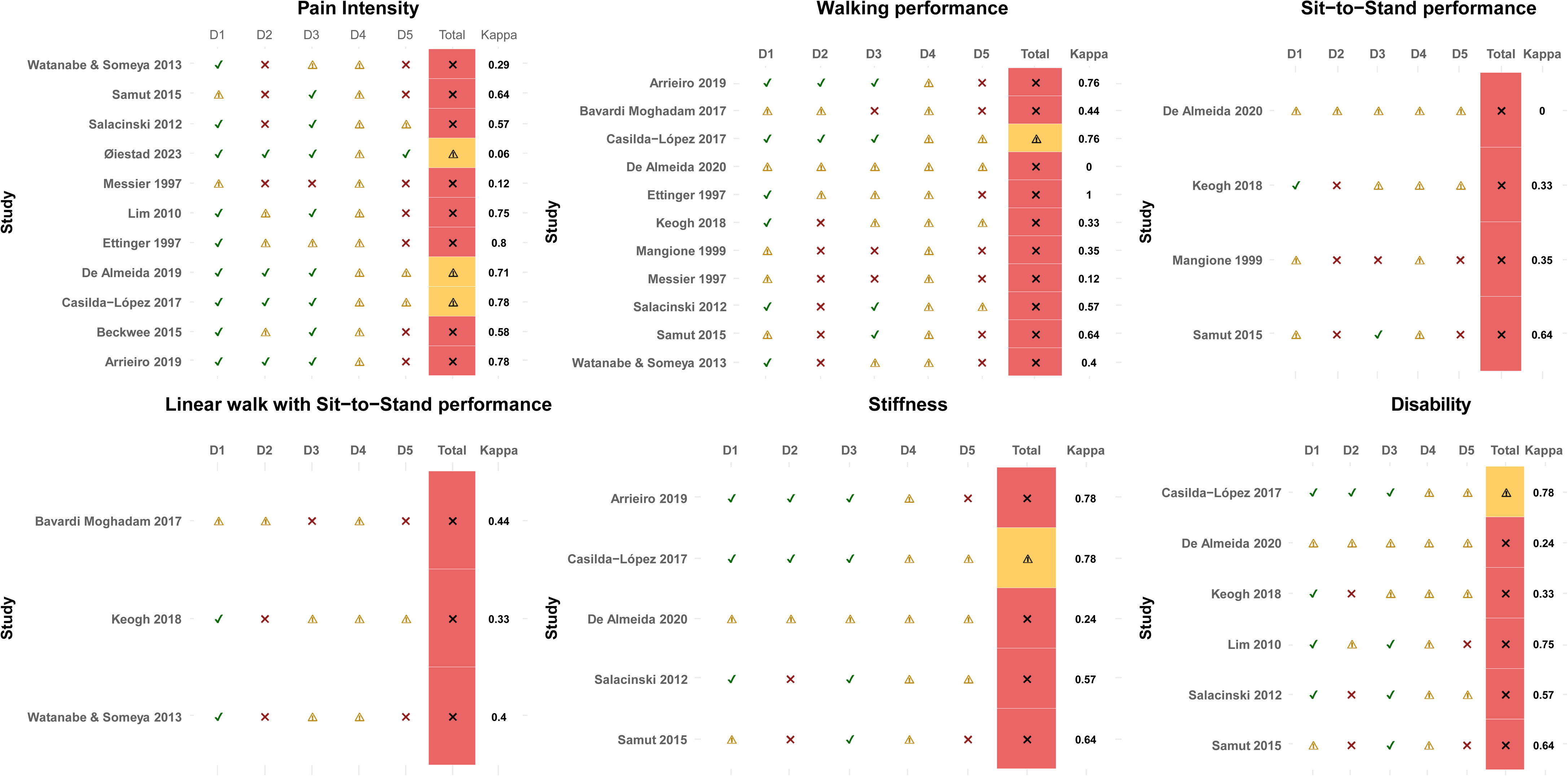

For walking performance, 10 studies presented “high risk” of bias (Arrieiro et al., 2019; Bavardi Moghadam & Shojaedin, 2017; de Almeida et al., 2020; Ettinger et al., 1997; Keogh et al., 2018; Mangione et al., 1999; Messier et al., 1997; Salacinski et al., 2012; Samut et al., 2015; Watanabe & Someya, 2013), while 1 study presented “some concerns” (Casilda-López et al., 2017). Inter-rater agreement ranged from from slight (κ = 0) to perfect (κ = 1), see Figures 3.

All studies evaluating sit-to-stand performance presented a “high risk” of bias (de Almeida et al., 2020; Keogh et al., 2018; Mangione et al., 1999; Samut et al., 2015). Inter-rater agreement for this outcome ranged from slight (κ = 0) to substantial (κ = 0.64), see Figures 3.

Similarly, all studies assessing combined sit-to-stand and walking performance were rated with “high risk” of bias (Bavardi Moghadam & Shojaedin, 2017; Keogh et al., 2018; Watanabe & Someya, 2013), with an inter-rater agreement ranging from from fair (κ = 0.33) to moderate (κ = 0.44), see Figures 3.

For the perceived stiffness, 3 studies showed “high risk” of bias (Arrieiro et al., 2019; de Almeida et al., 2020; Salacinski et al., 2012), while 1 presented “some concerns” (Casilda-López et al., 2017). Inter-rater agreement ranged from fair (κ = 0.24) to substantial (κ = 0.78).

Finally, regarding disability related to KOA, 4 studies were rated with a “high risk” of bias (de Almeida et al., 2020; Keogh et al., 2018; Lim et al., 2010; Salacinski et al., 2012), while 1 study presented “some concerns” (Casilda-López et al., 2017). Inter-rater agreement ranged from fair (κ = 0.33) to substantial (κ = 0.78), see Figure 3.

### 3.4. Network meta-analysis

Two meta-analyses were conducted: one focused on pain intensity, and the other on walking performance. Although 6 outcomes were initially considered, the remaining 4 could not be analyzed due to insufficient connectivity between studies in the network. Details on the eligibility process for including studies in the meta-analyses, data availability, instrument selection, and the procedures used to estimate means and SD are presented in Table 2.

#### 3.4.1. Pain intensity

Seven studies were included in the meta-analysis (Beckwée et al., 2015; de Almeida et al., 2019; Ettinger et al., 1997; Lim et al., 2010; Øiestad et al., 2023; Salacinski et al., 2012; Samut et al., 2015) examining the effects of different aerobic exercise intensities. These included light-to-moderate-to-vigorous (de Almeida et al., 2019), moderate (Salacinski et al., 2012), moderate-to-vigorous (Ettinger et al., 1997; Lim et al., 2010; Øiestad et al., 2023) and vigorous intensities (Beckwée et al., 2015; Samut et al., 2015). No direct comparisons between aerobic exercise intensities were available. Control groups included educational protocols (de Almeida et al., 2019; Ettinger et al., 1997), resistance exercise (Beckwée et al., 2015; de Almeida et al., 2019; Ettinger et al., 1997; Øiestad et al., 2023; Samut et al., 2015), no treatment (Salacinski et al., 2012), and multimodal exercise (resistance, aerobic and stretching) interventions (Lim et al., 2010). See Figure 4.

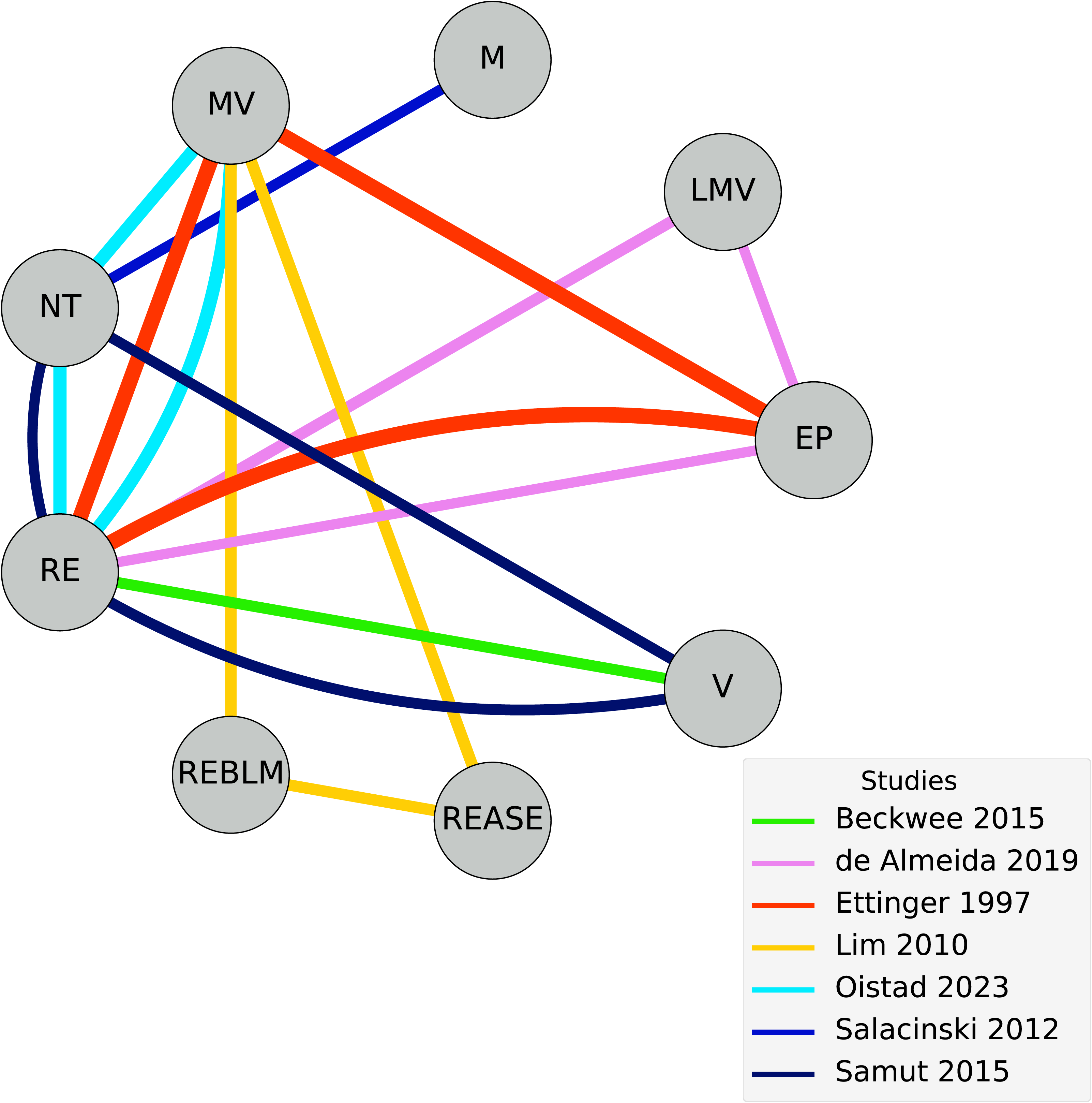

The two NMA models revealed non-significant and imprecise effects for all AE intensities, including light-to-moderate-to-vigorous (Model 1: g = □0.27; 95%CrI □3.19, 2.67; Model 2: g = □0.27; 95%CrI □3.19, 2.78), moderate (Model 1: g = □0.65; 95%CrI □4.09, 2.99; Model 2: g = □0.59; 95%CrI □7.07, 5.98), moderate-to-vigorous intensities (Model 1: g = □0.42; 95%CrI □2.49, 1.56; Model 2: g = □0.42; 95%CrI □2.48, 1.59), and vigorous (Model 1: g = 0; 95%CrI □2.19, 2.17; Model 2: g = 0; 95%CrI □2.17, 2.19) compared to “no treatment”, see Figure 5 and 6.

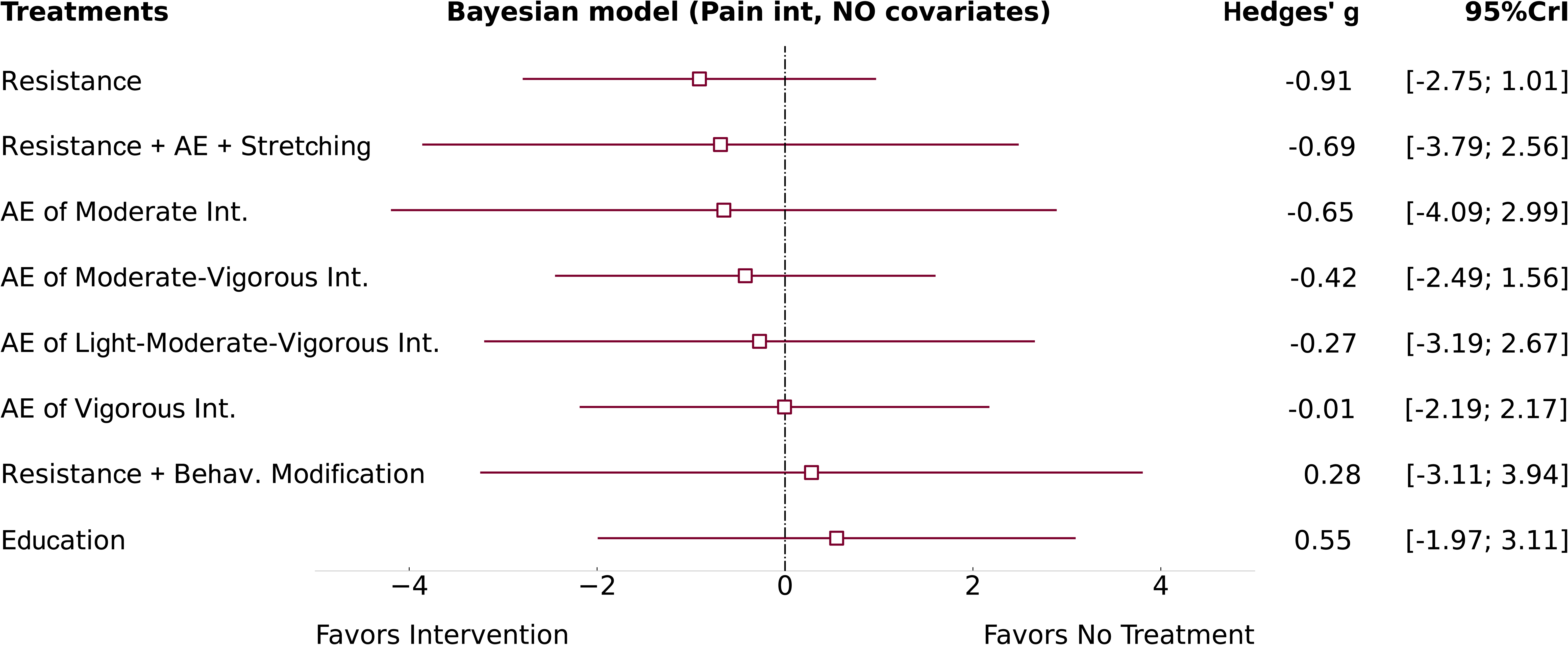

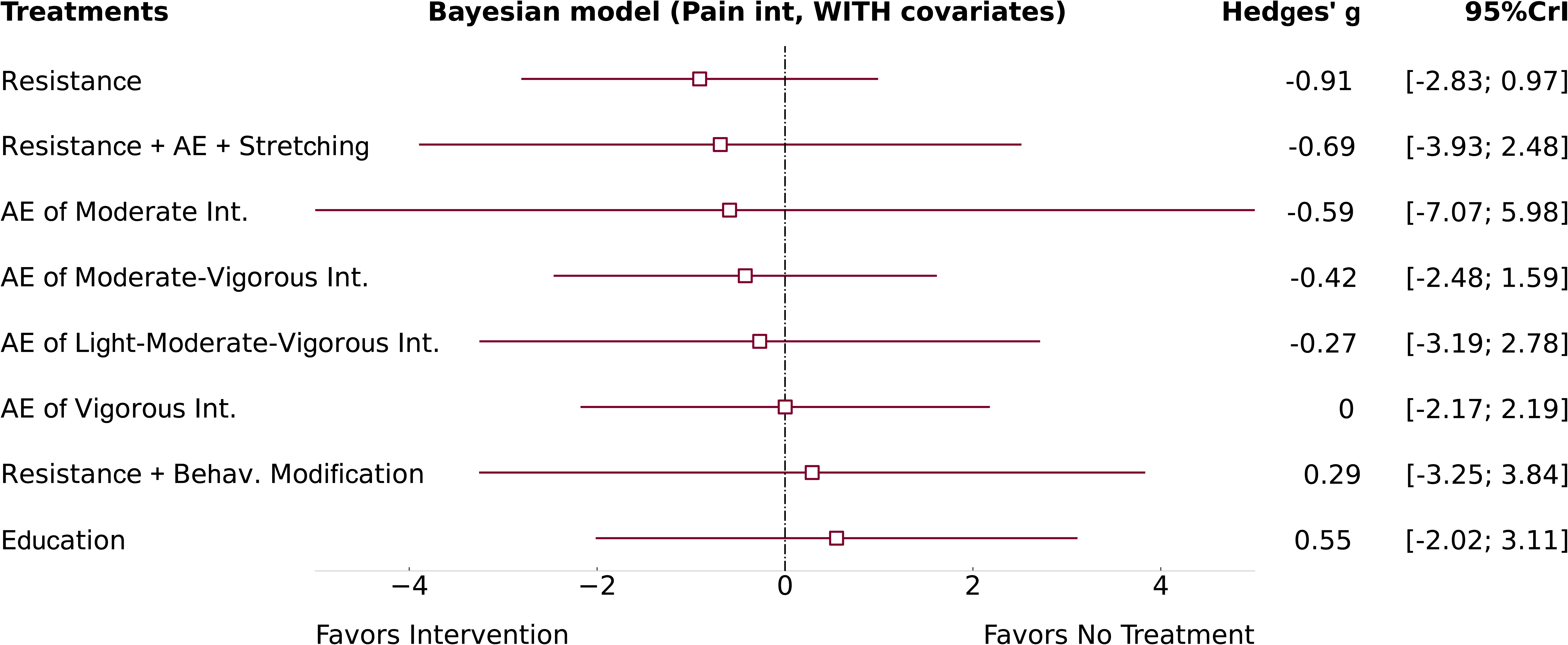

The addition of both covariates (weekly frequency and number of weeks) did not meaningfully adjust the precision of AE intensity estimates. In addition, neither weekly frequency (w_f_ = □0.15; 95%CrI □19.15, 18.28) nor number of treatment weeks (w_n_ = □0.09; 95%CrI □19.34, 19.30) showed a linear association with the observed effects.

Heterogeneity was large in both models: Model 1, τ = 1.40; 95%CrI 0.70, 2.26; and Model 2, τ = 1.40; 95%CrI 0.69, 2.29.

Comparisons between different intensities of aerobic exercise were based exclusively on indirect evidence, due to the absence of direct pairwise comparisons. All estimated effects from both models were non-significant, with 95%CrI exceedingly wide in both directions, including large effect sizes (g>0.7). This indicates a high degree of imprecision and limits the ability to draw reliable conclusions about the true effect of aerobic exercise intensities. See Table 3 and Figure 7.

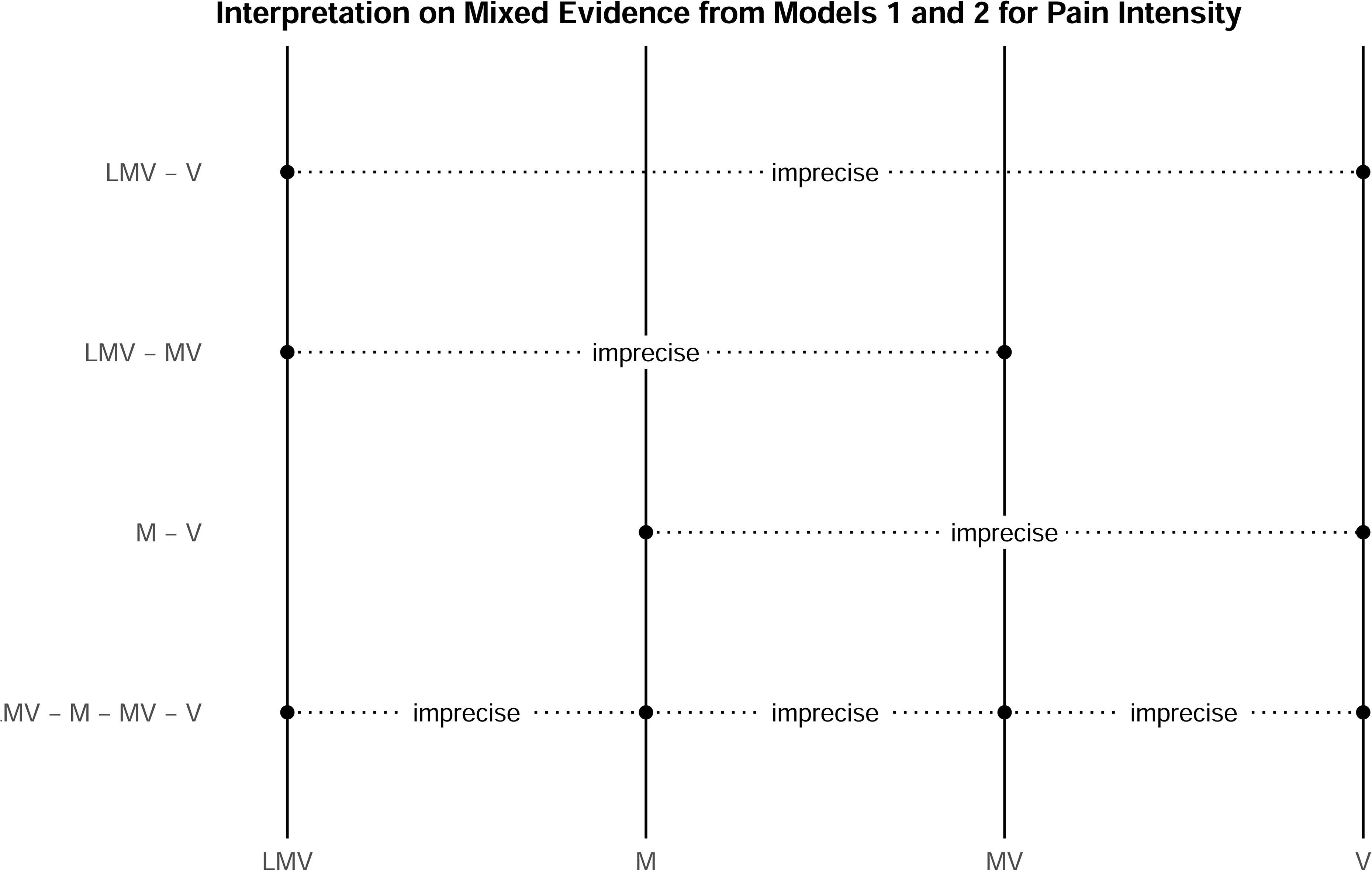

The preliminary GRADE assessment of the indirect network for both models identified “very serious” concerns regarding risk of bias, while concerns for indirectness were judged as “not serious”. Heterogeneity was rated as “very serious” limitation, whereas intransitivity was not detected and therefore judged as “not serious”. Publication bias across the indirect network ranged from “not serious” to “very serious” depending on the comparison, see Figures 8 and 9. For the mixed (NMA) estimates, imprecision was consistently judged as “very serious” across all comparisons. Consequently, the final GRADE evaluation indicated an overall certainty of evidence of “very low” for all comparisons of AE intensities in both models, see Table 3.

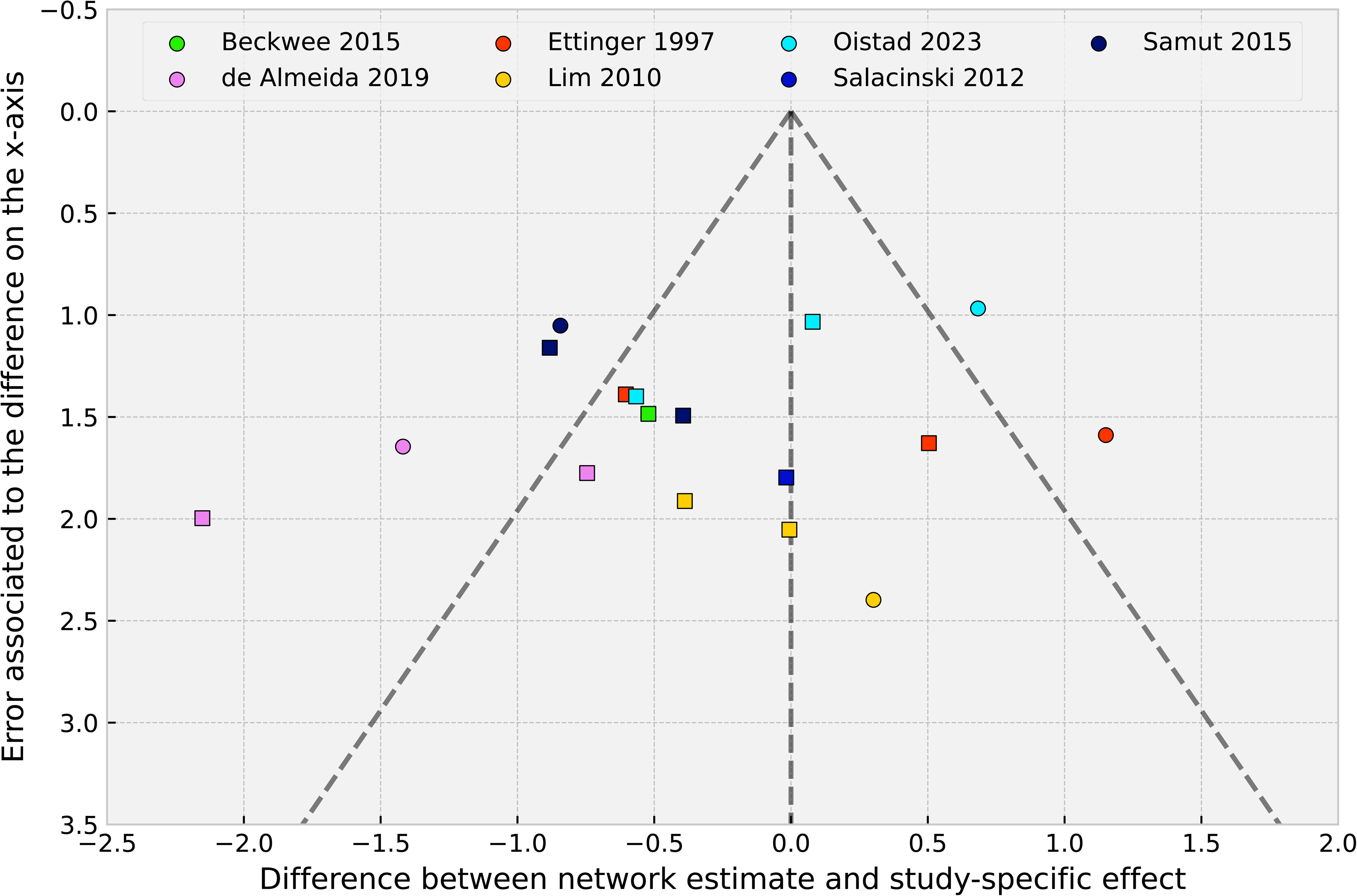

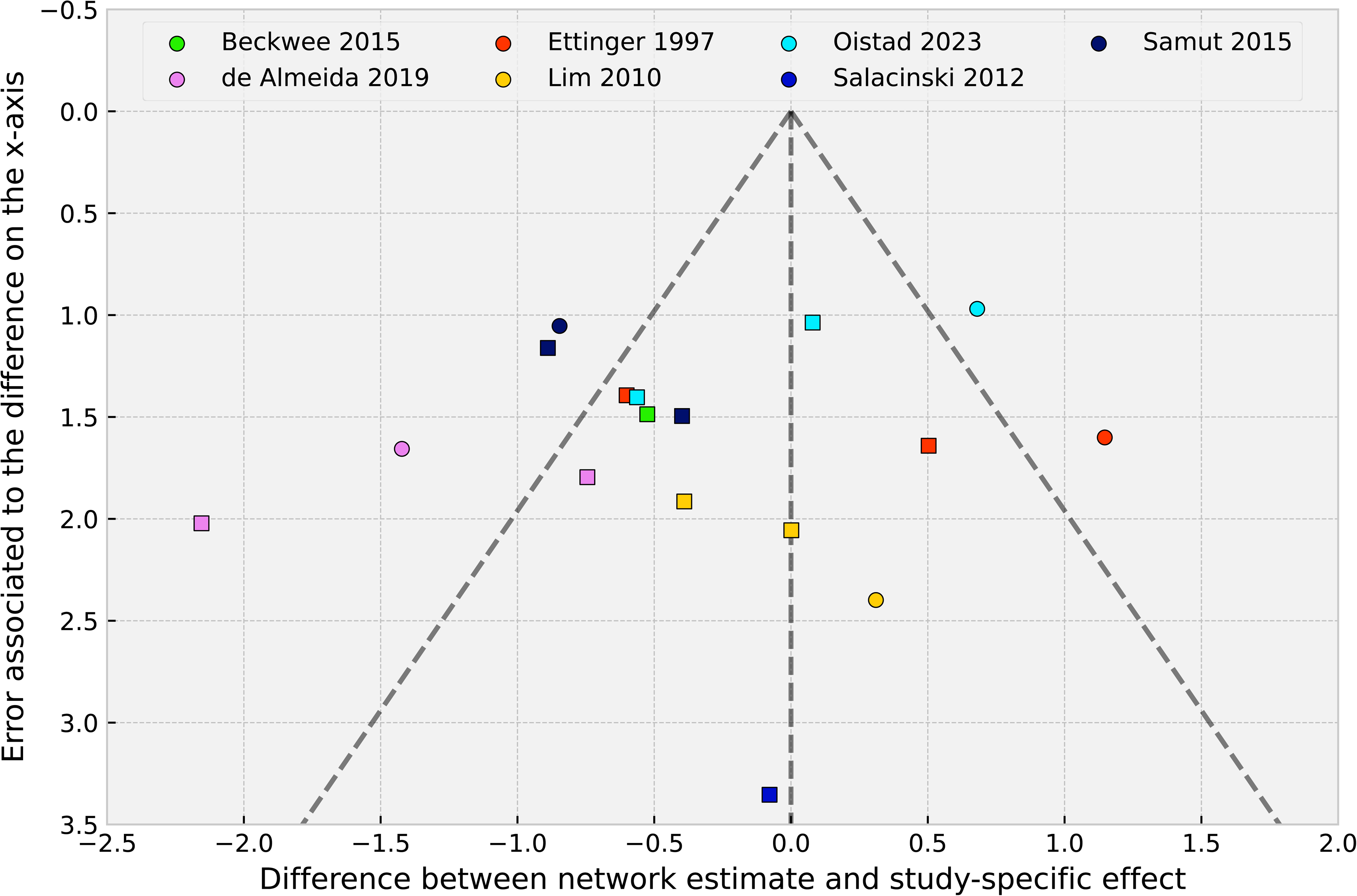

#### 3.4.2. Walking performance

Seven studies were included in the meta-analysis (Bavardi Moghadam & Shojaedin, 2017; de Almeida et al., 2020; Ettinger et al., 1997; Keogh et al., 2018; Mangione et al., 1999; Salacinski et al., 2012; Samut et al., 2015) examining the effects of different aerobic exercise intensities. These included light-to-moderate (Mangione et al., 1999), light-to-moderate-to-vigorous (de Almeida et al., 2020), moderate (Keogh et al., 2018; Salacinski et al., 2012), moderate-to-vigorous (Bavardi Moghadam & Shojaedin, 2017; Ettinger et al., 1997), and vigorous intensities (Keogh et al., 2018; Mangione et al., 1999; Samut et al., 2015). Two direct comparisons between intensities were available: light-to-moderate versus vigorous (Mangione et al., 1999) and moderate versus vigorous (Keogh et al., 2018).

Control groups included resistance exercise (Beckwée et al., 2015; de Almeida et al., 2020; Ettinger et al., 1997; Lim et al., 2010; Øiestad et al., 2023; Samut et al., 2015), educational protocols (de Almeida et al., 2020; Ettinger et al., 1997; Samut et al., 2015) and no treatment (Bavardi Moghadam & Shojaedin, 2017; Salacinski et al., 2012). See Figure 10.

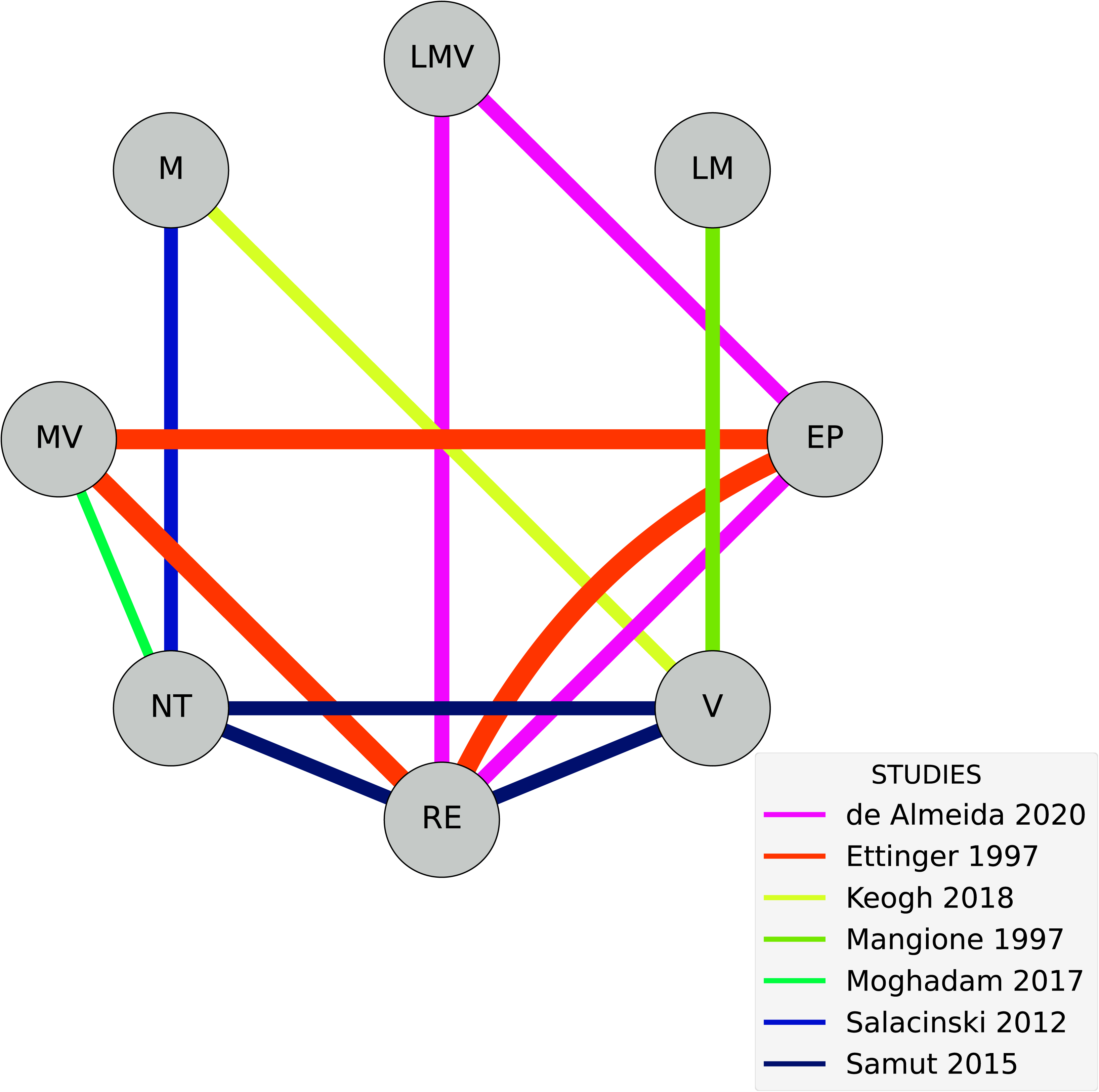

The two NMA models revealed non-significant and imprecise effects for all AE intensities, including light-to-moderate intensity (Model 1: g = 0.78; 95%CrI: –2.31, 3.88; Model 2: g = 0.33; 95%CrI: –1.34, 1.96), for light-to-moderate-to-vigorous intensity (Model 1: g = 1.22; 95%CrI: –1.12, 3.87; Model 2: g = 0.25; 95%CrI: –1.10, 1.62), for moderate aerobic exercise (Model 1: g = 0.89; 95%CrI: –2.31, 2.94; Model 2: g = 0.06; 95%CrI: –1.82, 1.91), for moderate-to-vigorous intensity (Model 1: g = 1.82; 95%CrI: –0.21, 4.21; Model 2: g = 0.45; 95%CrI: –0.86, 1.72), and vigorous intensity (Model 1: g = 0.91; 95%CrI: –0.63, 2.61; Model 2: g = 0.46; 95%CrI: –0.42, 1.32), compared to “no treatment”. See Figures 11 and 12.

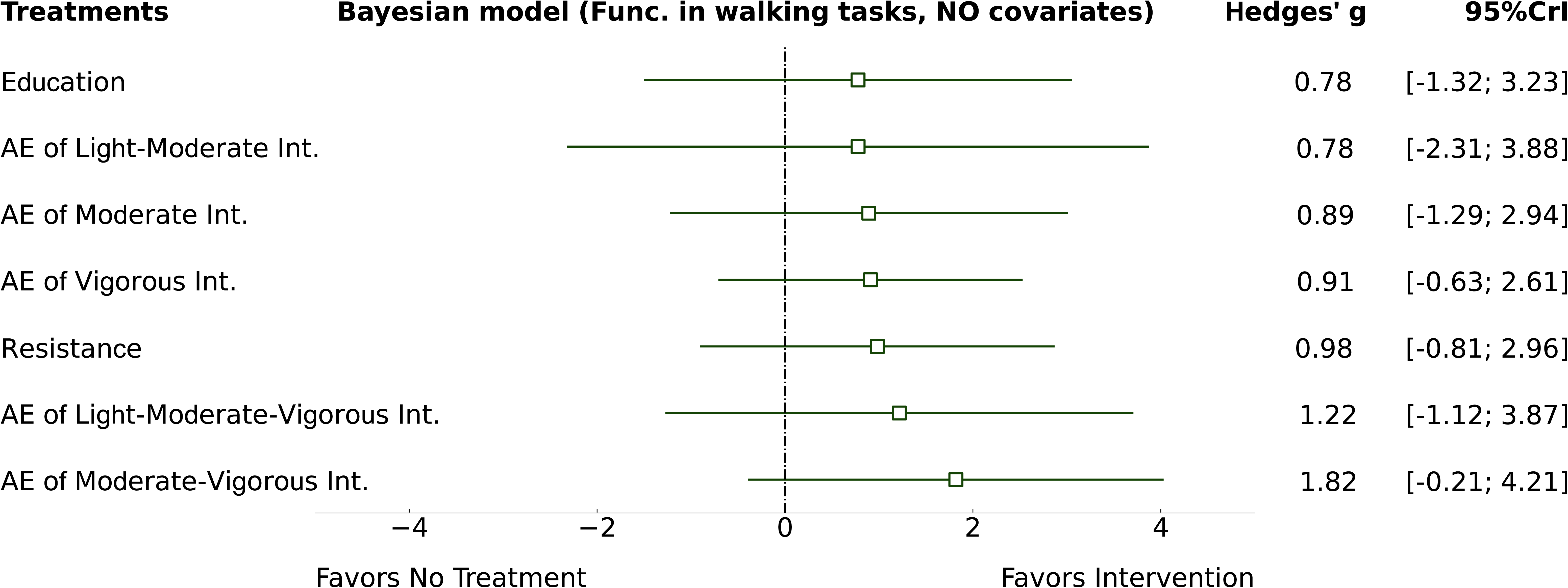

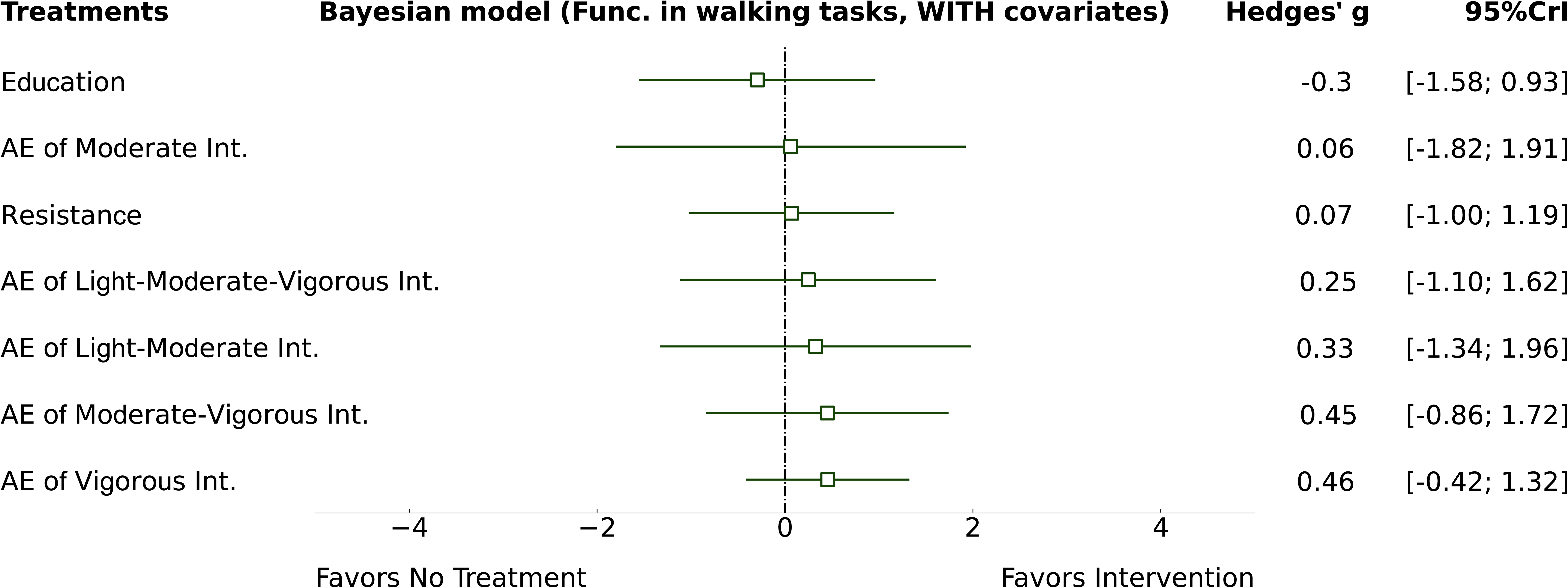

The inclusion of both covariates (weekly frequency and number of weeks) slightly improved the precision of AE intensity estimates. Weekly frequency (w_f_ = 2.68; 95%CrI 0.68, 4.62) showed a significant positive effect on walking function. However, the 95%CrI indicated an imprecise estimate, limiting the plausibility that can be drawn from this result. The upper bound of the 95% CrI suggests a value that is not plausible, while the lower bound already indicates a large effect (implying that each additional day would increase Hedges’ g by 0.68). The number of weeks (w_n_ = □0.35; 95%CrI □0.8, 0.1) showed a non-significant and imprecise association with the observed effects.

Heterogeneity was large in Model 1 (τ = 0.89; 95%CrI 0.17, 1.84), and decreased to a small in Model 2 (τ = 0.30; 95%CrI 0, 0.82).

Mixed evidence of pairwise comparatives between different intensities of aerobic exercise were obtained exclusively from indirect evidence because of the absence of direct evidence, except two of them: the comparative between light-to-moderate vs vigorous presented direct evidence without indirect evidence (Mangione et al., 1999). Finally, only the comparative between moderate vs vigorous (Keogh et al., 2018) presented both direct and indirect evidence.

Both models presented non-significant and imprecise results for pairwise comparisons, with 95%CrI, exceeding in both directions with large effect sizes (g>0.7), preventing drawing clear conclusions about the real difference between AE intensities, see Table 4 and Figure 13.

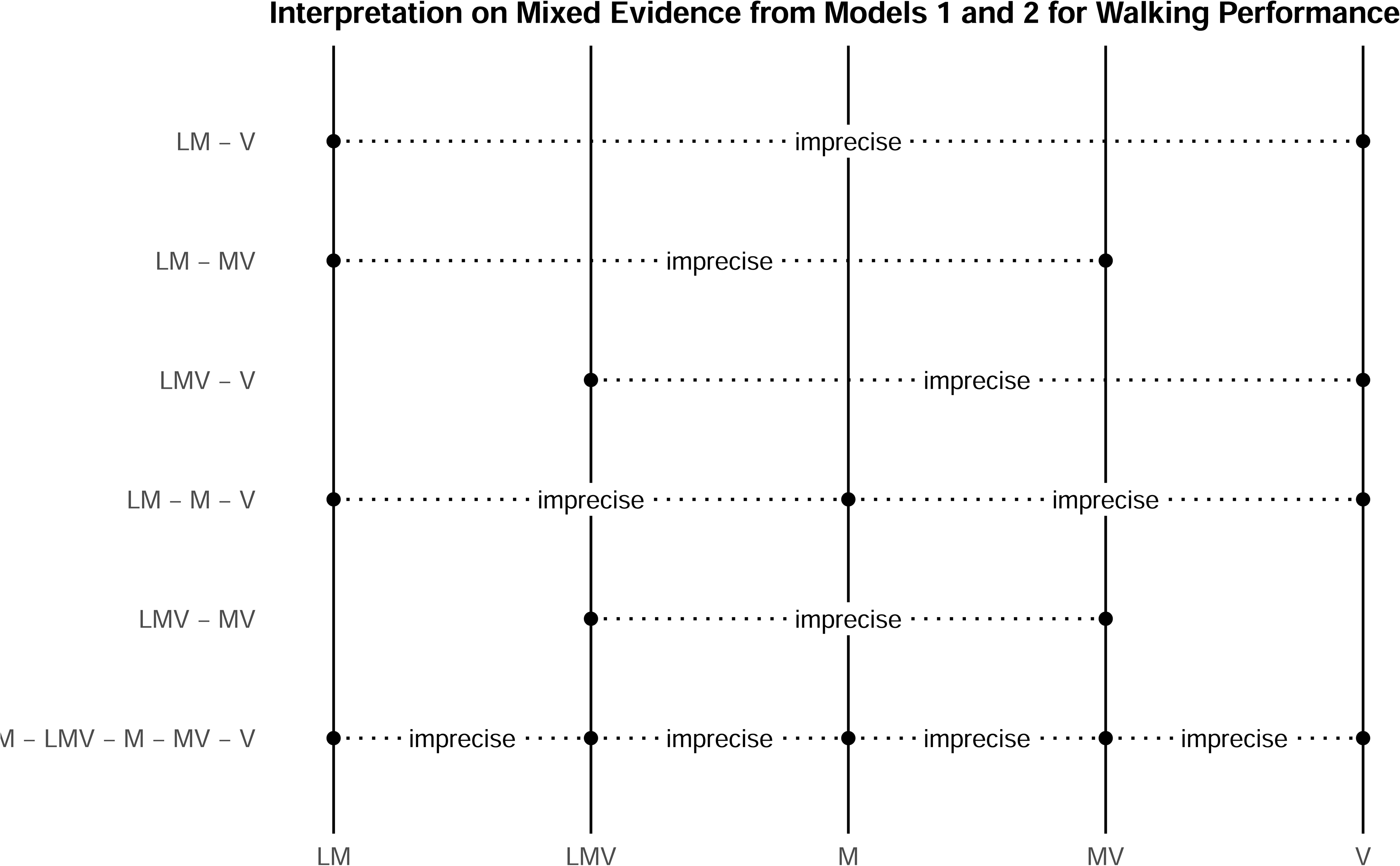

A preliminary GRADE assessment was conducted for direct networks. Risk of bias was judged as a “very serious” concern, while heterogeneity and publication bias could not be assessed because only one study contributed to each direct comparison, therefore, these domains were rated as “not serious” (see Figures 14 and 15, and Table 4).

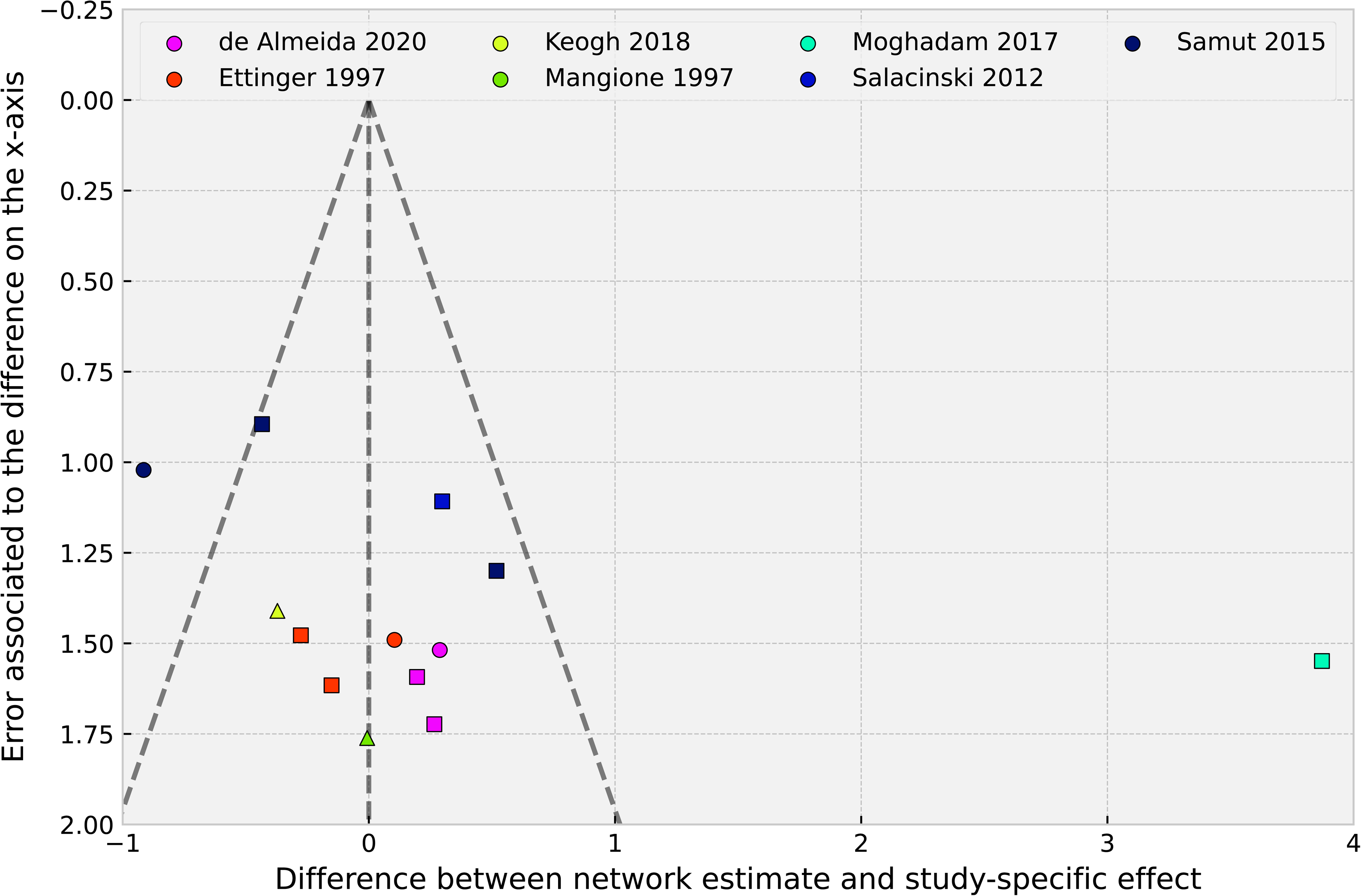

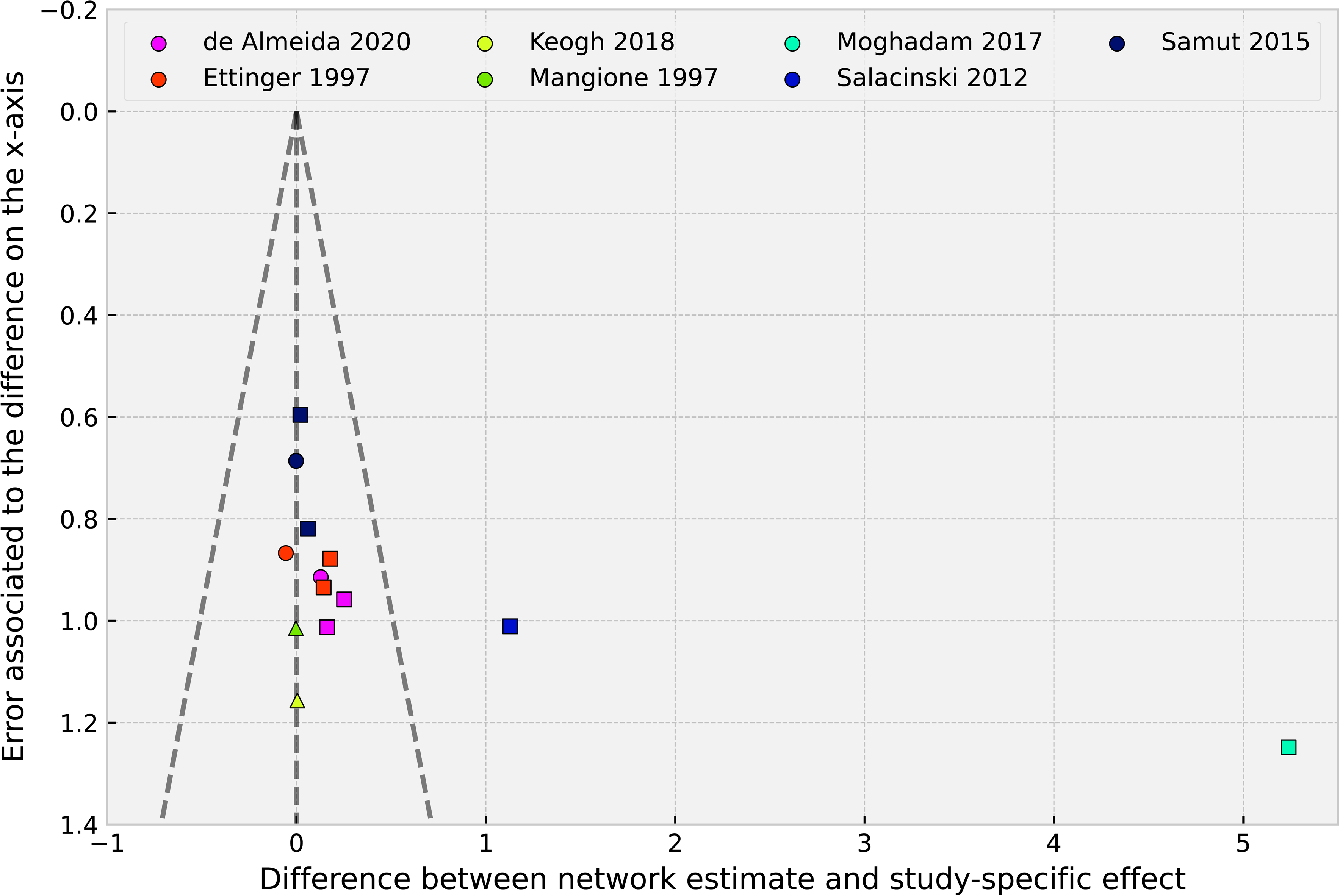

The preliminary GRADE evaluation of the indirect networks revealed “very serious” concerns for risk of bias, whereas indirectness was judged as “not serious”. Heterogeneity was rated as “very serious” across all comparisons in Model 1 and ranged from “not serious” to “very serious” in Model 2. Publication bias also ranged from “not serious” to “very serious” across comparisons in both models (Figures 14 and 15). Intransitivity was consistently judged as “not serious” in all comparisons.

For the mixed evidence provided “very serious” imprecision in all comparisons and models, with all comparisons judged with a “very low” certainty of evidence, see Table 3.

## 4. Discussion

In this review, we aimed to determine which intensity of aerobic exercise is most effective for patients with KOA for the following six outcomes: knee pain intensity, performance in walking tasks, performance in sit-to-stand, performance in combined sit-to-stand and walking tasks, perceived knee stiffness and disability related with KOA. For this purpose, it was proposed the creation of a NMA to estimate the effects of aerobic exercise intensities.

Fifteen studies (Arrieiro et al., 2019; Bavardi Moghadam & Shojaedin, 2017; Beckwée et al., 2015; Casilda-López et al., 2017; de Almeida et al., 2019, 2020; Ettinger et al., 1997; Keogh et al., 2018; Lim et al., 2010; Mangione et al., 1999; Messier et al., 1997; Øiestad et al., 2023; Salacinski et al., 2012; Samut et al., 2015; Watanabe & Someya, 2013) were finally included in this review. On the other hand, only two meta-analyses were developed based on the network availability, analysing pain intensity and performance in walking tasks.

No clear conclusions could be drawn from neither of the meta-analyses, as high risk of bias, and imprecise CrI were present. This imprecision was mainly derived from two factors: 1) lack of statistical power due to the low number of available studies in each aerobic exercise intensity category; and 2) due to the NMA evidence was exclusively derived, in the majority of pairwise comparisons, from indirect evidence, as no direct comparisons were present. Clearly stablishing differences between similar interventions, for example, stablishing the amount of difference between light, and light-to-moderate intensities is a difficult task, and in fact, it is plausible that these differences in outcome measures such as pain intensity may not be precise, mainly due to the multifactorial nature of pain. In the case of performance outcome measures, it would be expected that they increase with exercise intensity. However, the opposite case could also appear, as greater intensities may increase the amount of flare-ups in KOA symptomatology, increasing pain intensity and reducing patients’ functioning. The scope to answer these questions were limited through 2 factors: 1) the lack detailed reports in drop-out motives; and 2) the scarce methods for reducing the bias derived from not analysing drop-outs (such as data-imputation analyses). This was integrated as part of the evaluation in the ROB tool, in Domain n°2. Five of the seven studies included in the pain intensity NMA presented “some concerns” or “high risk” of bias (Beckwée et al., 2015; Ettinger et al., 1997; Lim et al., 2010; Salacinski et al., 2012; Samut et al., 2015) and all the studies in walking performance NMA presented “some concerns” or “high risk” in Domain n°2.

To our knowledge our study is the first to compare different aerobic exercise intensities in patients with osteoarthritis of the knee. In a previous study, but including strength exercise, Regnaux et al., (2015) compared high intensity exercise with low intensity exercise and observed a statistically significant difference in favor of high intensity exercise in terms of pain and function, but concluded that they are not clinically relevant. Some studies have also compared the efficacy of strength training to aerobic exercise, in terms of pain and function, and found neither to be superior to the other (Ceballos-Laita et al., 2023).

Although aerobic exercise, as an isolated intervention, has been considered a potentially effective treatment for patients with KOA, this conclusion warrants careful interpretation in light of several limitations that may affect the robustness and generalizability of available evidence. These limitations are consistently observed across most of the systematic reviews with meta-analyses published to date, largely due to the methodological criteria commonly employed in their study selection and synthesis processes. Such limitations include: 1) the inclusion of patients with other comorbidities, such as hip OA, within KOA study populations (Goh et al., 2019; Luo et al., 2025; Regnaux et al., 2015; Uthman et al., 2014) ; 2) the absence of standardized criteria for defining and parametrizing aerobic exercise (Goh et al., 2019; Juhl et al., 2014; Luo et al., 2025); 3) the frequent combination of aerobic exercise with other interventions, such as physiotherapy, usual care, lifestyle recommendations, educational programs, self-management programs, balance or flexibility exercised or others (Goh et al., 2019; Juhl et al., 2014; Luo et al., 2025); 4) controversial risk of bias assessments (Luo et al., 2025); 5) low quality of the available evidence according to the GRADE approach (Regnaux et al., 2015); 6) large credible intervals observed in the results, which increase imprecision and make it difficult to draw a clear conclusion for certain comparisons and outcomes (Juhl et al., 2014).

The present review aimed to address these concerns of extrapolability, employing rigorous criteria of study selection. However, a low number of studies were finally identified. This poses the question whether aerobic exercise still present a considerable high-quality evidence for supporting its results, as considered in previous studies (Fransen et al., 2015).

Exercise, and specifically aerobic modality, presents several plausible justifications that may indicate an improvement in symptoms related to KOA. These include the effects of aerobic exercise in pain modulation central mechanisms; its role in the reduction of systemic an articular pro-inflammatory markers, which are involved in the pathogenesis of KOA; its effects on the improvement of physical functioning in KOA patients, therefore enhancing their functional level regardless of their symptomatic state; a collateral interventor on the psychological state of chronic pain patients. All these mechanisms support a multifactorial model of action of aerobic exercise.

From a neurophysiological perspective, aerobic exercise may influence central mechanisms of pain modulation. Studies have shown its ability to stimulate the release of neurotransmitters such as serotonin, which play a key role in descending pain inhibition, as well as to promote neuroplasticity through the upregulation of brain-derived neurotrophic factor and other neurotrophic agents involved in neural adaptation and pain processing (Al-Sharman et al., 2019; Puts et al., 2023).

Regarding inflammation, aerobic exercise has been associated with a reduction in systemic and articular pro-inflammatory mediators (Knights et al., 2022), such as cytokines, which are known to contribute to the pathogenesis and progression of KOA (Primorac et al., 2020). While some clinical studies have reported decreases in these inflammatory markers following exercise interventions (Puts et al., 2023). Supporting these findings, preclinical studies rat models with induced-KOA have shown that aerobic exercise can decrease the levels of IL-1β, caspase-3, and MMP-13, helping to prevent cartilage breakdown (Assis et al., 2016). Other studies in similar models have also reported reductions in inflammatory markers following aerobic exercise, suggesting its potential role in modulating inflammatory processes associated with KOA (Martins et al., 2019). These effects may even contribute to slowing or preventing the progression of the disease (Rios et al., 2019).

Functionally, aerobic exercise (although with other interventions) has demonstrated beneficial effects on physical capacity, potentially enhancing gait performance, endurance, and general mobility in patients with KOA (Goh et al., 2019). These improvements in function may occur independently of changes in pain, contributing to a better overall level of activity. This supports the idea that exercise may have intrinsic functional benefits beyond symptom relief.

In addition, aerobic exercise may exert positive effects on psychological variables commonly associated with chronic pain, including stress, anxiety, and sleep disturbances. These psychosocial factors can perpetuate or exacerbate pain perception and disability, and their improvement may contribute indirectly to better clinical outcomes (Parmelee et al., 2015; Xu et al., 2024).

Taken together, these hypotheses support the potential of aerobic exercise as a promising intervention for KOA. Despite current limitations regarding its isolated effectiveness and the uncertainty around optimal intensity, existing evidence points to beneficial effects.

### 4.1. Limitations

This study presents several limitations. Firstly, there is a lack of studies that meet the inclusion criteria, due to heterogeneity in population characteristics, lack of detailed exercise prescription parameters and scarce studies including isolated aerobic exercise. The low number of studies included in the analysis reduces the statistical power to detect potential differences between different intensities of aerobic exercise. Secondly, there is a shortage of studies directly comparing different intensities of aerobic exercise. Most direct comparisons are found between different exercise modalities, or between aerobic exercise and usual care interventions, thus comparisons between aerobic exercise intensities were primarily indirect. The third limitation is the high risk of bias in the included studies.

### 4.2. Clinical implications

Aerobic exercise seems to be a promising treatment option for patients with KOA. However, there is a lack of available literature analyzing exclusively KOA patients, with isolated aerobic exercise treatments. Given the present results, aerobic exercise could be beneficial for this population, however, we lack certainty whether this modality, or its intensity variation may modulate pain intensity or function in KOA. Given this information, it is advisable for clinicians to adjust the exercise intensity based on the patient’s characteristics, preferences, and clinical response to exercise. Clinicians should always consider combining aerobic exercise with other interventions (self-management, physiotherapy treatment, educational programs or medication control), to warrant the effectiveness of the therapy protocol. Implementing a medium to long-term aerobic exercise program in these patients is likely beneficial due to its positive effects on commonly associated comorbidities such as overweight, hypertension, insulin tolerance, or metabolic syndrome.

## 5. Conclusions

Very low certainty of evidence limits drawing precise conclusions from the effects of aerobic exercise intensity in pain severity and performance in walking tasks. Comparatives showed concerning risk of bias, heterogeneity, certain publication bias, and significant imprecision of credible intervals in the pooled results. More studies with higher quality will be needed to determine which intensity of aerobic exercise is most effective for these patients.

## Supporting information

T1 Summary

T2 Data availability

T3 GREADE Pain

T4 GRADE Walking

## Data Availability

All data produced in the present work are contained in the manuscript

## 7. Author contributions

LCD, JPF, and ICG: investigation, data curation, visualization, writing – original draft. PC: conceptualization, data curation, formal analysis, funding acquisition, methodology, software, validation, visualization, writing – original draft, writing – review and editing. CDCL: validation, writing – original draft preparation, writing – reviewing and editing. JFM: investigation, conceptualization, data curation, visualization, formal analysis, methodology, supervision, validation, visualization, writing - original draft, writing – review and editing.

