## Supplementary Material for "How intense is effective? Exploring aerobic exercise intensity for knee osteoarthritis through a Bayesian network meta-analysis"

- Supplementary Material 1. Summary table of search engines, databases and search equations.
- Supplementary Material 2. Methods of Estimating Intensity of Cardiorespiratory Exercise by the ACSM 11^th^ Edition.
- Supplementary Material 3. Computation of Hedges’ g and its exact variance.
- Supplementary Material 4. Python syntax for computing Bayesian NMA, pairwise comparisons and funnel plots.

Supplementary Material 1. Summary table of search engines, databases and search equations.

| **Search Engine** | **Databases** | **Searches (nº)** | **Equation** | **Date** | **Registries (n)** |
| --- | --- | --- | --- | --- | --- |
| PubMed | MEDLINE | Nº1 | (("osteoarthritis, knee"[MeSH Terms] OR "gonarthrosis"[Title/Abstract] OR (("Knee"[Title/Abstract] OR "Tibiofemoral"[Title/Abstract] OR "Femorotibial"[Title/Abstract] OR “Knee”[MeSH Terms] OR "Knee Joint"[MeSH Terms]) AND ("Osteoarthri*"[Title/Abstract] OR "Arthrosi*"[Title/Abstract] OR "Osteoarthrosi*"[Title/Abstract] OR "Osteoarthritis"[MeSH Terms] OR (("Cartilage"[Title/Abstract] OR "Chondral"[Title/Abstract]) AND ("Defect*"[Title/Abstract] OR "Loss"[Title/Abstract] OR "Volume"[Title/Abstract] OR "Density"[Title/Abstract] OR "Thickness"[Title/Abstract] OR "Structure"[Title/Abstract] OR "Infrastructure"[Title/Abstract] OR "Surface"[Title/Abstract] OR "Degenerat*"[Title/Abstract] OR "Deform*"[Title/Abstract]))))) AND ("Circuit-Based Exercise"[MeSH Terms] OR "Endurance Training"[MeSH Terms] OR "High-Intensity Interval Training"[MeSH Terms] OR "Running"[MeSH Terms] OR "Swimming"[MeSH Terms] OR "Walking"[MeSH Terms] OR "Running"[Title/Abstract] OR "Run"[Title/Abstract] OR "Jogging"[Title/Abstract] OR "Walk*"[Title/Abstract] OR "Cycling"[Title/Abstract] OR "Swimming"[Title/Abstract] OR (("Aerobic"[Title/Abstract] OR "Endurance"[Title/Abstract] OR "Cardiovascular"[Title/Abstract] OR "Cardiopulmonar*"[Title/Abstract] OR "Cardiorespiratory"[Title/Abstract] OR "Metabolic*"[Title/Abstract] OR "Circuit"[Title/Abstract] OR "Circuit-based"[Title/Abstract] OR "Interval*"[Title/Abstract]) AND ("Exercis*"[Title/Abstract] OR "Train*"[Title/Abstract] OR "Workout*"[Title/Abstract] OR "Practice"[Title/Abstract] OR "Activit*"[Title/Abstract] OR "Conditioning"[Title/Abstract]))) AND ("Trial"[Title/Abstract] OR "randomized controlled trial"[Publication Type] OR "Randomized Controlled Trials as Topic"[MeSH Terms])) NOT ("Review"[Title] OR "Systematic review"[Title] OR "Meta analysis"[Title] OR "Metaanal*"[Title] OR "Meta anal*"[Title] OR "Review"[Publication Type] OR "Systematic review"[Publication Type] OR "Review Literature as Topic"[MeSH Terms] OR "Meta analysis"[Publication Type] OR "Meta-Analysis as Topic"[MeSH Terms]) | 26^th^ Dec, 2023 | 1344 |
| EBSCO | CINAHL Complete, SPORTDiscuss with Full Text, EBSCO eClassics Collection (EBSCOhost), OpenDissertations | Nº1 | ((AB "Knee osteoarthritis" OR TI "Knee osteoarthritis" OR AB "Gonarthrosis" OR TI "Gonarthrosis" OR ((AB "Knee" OR TI "Knee" OR AB "Tibiofemoral" OR TI "Tibiofemoral" OR AB "Femorotibial" OR TI "Femorotibial") AND (AB "Osteoarthri*" OR TI "Osteoarthri*" OR AB "Arthrosi*" OR TI "Arthrosi*" OR AB "Osteoarthrosi*" OR TI "Osteoarthrosi*" OR ((AB "Cartilage" OR TI "Cartilage" OR AB "Chondral" OR TI "Chondral") AND (AB "Defect*" OR TI "Defect*" OR AB "Loss" OR TI "Loss" OR AB "Volume" OR TI "Volume" OR AB "Density" OR TI "Density" OR AB "Thickness" OR TI "Thickness" OR AB "Structure" OR TI "Structure" OR AB "Infrastructure" OR TI "Infrastructure" OR AB "Surface" OR TI "Surface" OR AB "Degenerat*" OR TI "Degenerat*" OR AB "Deform*" OR TI "Deform*"))))) AND (AB "Circuit-Based Exercise" OR TI "Circuit-Based Exercise" OR AB "Endurance Training" OR TI "Endurance Training" OR AB "High-Intensity Interval Training" OR TI "High-Intensity Interval Training" OR AB "Running" OR TI "Running" OR AB "Swimming" OR TI "Swimming" OR AB "Walking" OR TI "Walking" OR AB "Running" OR TI "Running" OR AB "Run" OR TI "Run" OR AB "Jogging" OR TI "Jogging" OR AB "Walk*" OR TI "Walk*" OR AB "Cycling" OR TI "Cycling" OR AB "Swimming" OR TI "Swimming" OR ((AB "Aerobic" OR TI "Aerobic" OR AB "Endurance" OR TI "Endurance" OR AB "Cardiovascular" OR TI "Cardiovascular" OR AB "Cardiopulmonar*" OR TI "Cardiopulmonar*" OR AB "Cardiorespiratory" OR TI "Cardiorespiratory" OR AB "Metabolic*" OR TI "Metabolic*" OR AB "Circuit" OR TI "Circuit" OR AB "Circuit-based" OR TI "Circuit-based" OR AB "Interval*" OR TI "Interval*") AND (AB "Exercis*" OR TI "Exercis*" OR AB "Train*" OR TI "Train*" OR AB "Workout*" OR TI "Workout*" OR AB "Practice" OR TI "Practice" OR AB "Activit*" OR TI "Activit*" OR AB "Conditioning" OR TI "Conditioning"))) AND (AB "Trial" OR TI "Trial" OR AB "randomized controlled trial" OR TI "randomized controlled trial")) NOT (TI "Review" OR TI "Systematic review" OR TI "Meta analysis" OR TI "Metaanal*" OR TI "Meta anal*") | 26^th^ Dec, 2023 | 593 |
| Web of Science | Web of Science Core Collection, Current Contents Connect, Derwent Innovations Index, KCI-Korean Journal Database, ProQuest ™ Dissertations & Theses Citation Index, SciELO Citation Index | Nº1 | ((TS=("Knee osteoarthritis") OR TS=("Gonarthrosis") OR ((TS=("Knee") OR TS=("Tibiofemoral") OR TS=("Femorotibial")) AND (TS=("Osteoarthri*") OR TS=("Arthrosi*") OR TS=("Osteoarthrosi*") OR ((TS=("Cartilage") OR TS=("Chondral")) AND (TS=("Defect*") OR TS=("Loss") OR TS=("Volume") OR TS=("Density") OR TS=("Thickness") OR TS=("Structure") OR TS=("Infrastructure") OR TS=("Surface") OR TS=("Degenerat*") OR TS=("Deform*")))))) AND (TS=("Circuit-Based Exercise") OR TS=("Endurance Training") OR TS=("High-Intensity Interval Training") OR TS=("Running") OR TS=("Swimming") OR TS=("Walking") OR TS=("Running") OR TS=("Run") OR TS=("Jogging") OR TS=("Walk*") OR TS=("Cycling") OR TS=("Swimming") OR ((TS=("Aerobic") OR TS=("Endurance") OR TS=("Cardiovascular") OR TS=("Cardiopulmonar*") OR TS=("Cardiorespiratory") OR TS=("Metabolic*") OR TS=("Circuit") OR TS=("Circuit-based") OR TS=("Interval*")) AND (TS=("Exercis*") OR TS=("Train*") OR TS=("Workout*") OR TS=("Practice") OR TS=("Activit*") OR TS=("Conditioning")))) AND (TS=("Trial") OR TS=("randomized controlled trial"))) NOT (TI=("Review") OR TI=("Systematic review") OR TI=("Meta analysis") OR TI=("Metaanal*") OR TI=("Meta anal*")) | 26^th^ Dec, 2023 | 1960 |
| ScienceDirect | ScienceDirect | Nº1 | Title, abstract, keywords: (("Gonarthrosis" OR ("Knee" AND ("Osteoarthritis" OR "Arthrosis" OR "Osteoarthrosis"))) AND "Aerobic" AND ("Exercise" OR "Training") AND “Trial”)  Title: NOT ("Review" OR "Systematic review" OR "Meta analysis") | 26^th^ Dec, 2023 | 12 |
|  |  | Nº2 | Title, abstract, keywords: (("Gonarthrosis" OR ("Knee" AND ("Osteoarthritis" OR "Arthrosis" OR "Osteoarthrosis"))) AND "Endurance" AND ("Exercise" OR "Training") AND “Trial”)  Title: NOT ("Review" OR "Systematic review" OR "Meta analysis") | 26^th^ Dec, 2023 | 3 |
|  |  | Nº3 | Title, abstract, keywords: (("Gonarthrosis" OR ("Knee" AND ("Osteoarthritis" OR "Arthrosis" OR "Osteoarthrosis"))) AND "Cardiovascular" AND ("Exercise" OR "Training") AND “Trial”)  Title: NOT ("Review" OR "Systematic review" OR "Meta analysis") | 26^th^ Dec, 2023 | 4 |
|  |  | Nº4 | Title, abstract, keywords: (("Gonarthrosis" OR ("Knee" AND ("Osteoarthritis" OR "Arthrosis" OR "Osteoarthrosis"))) AND "Cardiopulmonar" AND ("Exercise" OR "Training") AND “Trial”)  Title: NOT ("Review" OR "Systematic review" OR "Meta analysis") | 26^th^ Dec, 2023 | 0 |
|  |  | Nº5 | Title, abstract, keywords: (("Gonarthrosis" OR ("Knee" AND ("Osteoarthritis" OR "Arthrosis" OR "Osteoarthrosis"))) AND "Cardiorespiratory" AND ("Exercise" OR "Training") AND “Trial”)  Title: NOT ("Review" OR "Systematic review" OR "Meta analysis") | 26^th^ Dec, 2023 | 3 |
|  |  | Nº6 | Title, abstract, keywords: (("Gonarthrosis" OR ("Knee" AND ("Osteoarthritis" OR "Arthrosis" OR "Osteoarthrosis"))) AND "Metabolic" AND ("Exercise" OR "Training") AND “Trial”)  Title: NOT ("Review" OR "Systematic review" OR "Meta analysis") | 26^th^ Dec, 2023 | 4 |
|  |  | Nº7 | Title, abstract, keywords: (("Gonarthrosis" OR ("Knee" AND ("Osteoarthritis" OR "Arthrosis" OR "Osteoarthrosis"))) AND "Circuit" AND ("Exercise" OR "Training") AND “Trial”)  Title: NOT ("Review" OR "Systematic review" OR "Meta analysis") | 26^th^ Dec, 2023 | 2 |
|  |  | Nº8 | Title, abstract, keywords: (("Gonarthrosis" OR ("Knee" AND ("Osteoarthritis" OR "Arthrosis" OR "Osteoarthrosis"))) AND "Interval" AND ("Exercise" OR "Training") AND “Trial”)  Title: NOT ("Review" OR "Systematic review" OR "Meta analysis") | 26^th^ Dec, 2023 | 40 |
| Scopus | Scopus | Nº1 | ((TITLE-ABS ( "Knee osteoarthritis" ) OR TITLE-ABS ( "Gonarthrosis" ) OR ((TITLE-ABS ( "Knee" ) OR TITLE-ABS ( "Tibiofemoral" ) OR TITLE-ABS ( "Femorotibial" )) AND (TITLE-ABS ( "Osteoarthri*" ) OR TITLE-ABS ( "Arthrosi*" ) OR TITLE-ABS ( "Osteoarthrosi*" ) OR ((TITLE-ABS ( "Cartilage" ) OR TITLE-ABS ( "Chondral" )) AND (TITLE-ABS ( "Defect*" ) OR TITLE-ABS ( "Loss" ) OR TITLE-ABS ( "Volume" ) OR TITLE-ABS ( "Density" ) OR TITLE-ABS ( "Thickness" ) OR TITLE-ABS ( "Structure" ) OR TITLE-ABS ( "Infrastructure" ) OR TITLE-ABS ( "Surface" ) OR TITLE-ABS ( "Degenerat*" ) OR TITLE-ABS ( "Deform*" )))))) AND (TITLE-ABS ( "Circuit-Based Exercise" ) OR TITLE-ABS ( "Endurance Training" ) OR TITLE-ABS ( "High-Intensity Interval Training" ) OR TITLE-ABS ( "Running" ) OR TITLE-ABS ( "Swimming" ) OR TITLE-ABS ( "Walking" ) OR TITLE-ABS ( "Running" ) OR TITLE-ABS ( "Run" ) OR TITLE-ABS ( "Jogging" ) OR TITLE-ABS ( "Walk*" ) OR TITLE-ABS ( "Cycling" ) OR TITLE-ABS ( "Swimming" ) OR ((TITLE-ABS ( "Aerobic" ) OR TITLE-ABS ( "Endurance" ) OR TITLE-ABS ( "Cardiovascular" ) OR TITLE-ABS ( "Cardiopulmonar*" ) OR TITLE-ABS ( "Cardiorespiratory" ) OR TITLE-ABS ( "Metabolic*" ) OR TITLE-ABS ( "Circuit" ) OR TITLE-ABS ( "Circuit-based" ) OR TITLE-ABS ( "Interval*" )) AND (TITLE-ABS ( "Exercis*" ) OR TITLE-ABS ( "Train*" ) OR TITLE-ABS ( "Workout*" ) OR TITLE-ABS ( "Practice" ) OR TITLE-ABS ( "Activit*" ) OR TITLE-ABS ( "Conditioning" )))) AND (TITLE-ABS ( "Trial" ) OR TITLE-ABS ( "randomized controlled trial" ))) AND NOT (TITLE ( "Review" ) OR TITLE ( "Systematic review" ) OR TITLE ( "Meta analysis" ) OR TITLE ( "Metaanal*" ) OR TITLE ( "Meta anal*" )) | 26^th^ Dec, 2023 | 1186 |
| SciELO | SciELO | Nº1 | ((ab:(“Knee osteoarthritis”)) OR (ti:(“Knee osteoarthritis”)) OR (ab:(“Gonarthrosis”)) OR (ti:(“Gonarthrosis”)) OR (((ab:(“Knee”)) OR (ti:(“Knee”))) AND ((ab:(“Osteoarthritis”)) OR (ti:(“Osteoarthritis”)) OR (ab:(“Arthrosis”)) OR (ti:(“Arthrosis”)) OR (ab:(“Osteoarthrosis”)) OR (ti:(“Osteoarthrosis”))))) AND ((ab:(“Running”)) OR (ti:(“Running”)) OR (ab:(“Swimming”)) OR (ti:(“Swimming”)) OR (ab:(“Walking”)) OR (ti:(“Walking”)) OR (ab:(“Running”)) OR (ti:(“Running”)) OR (ab:(“Run”)) OR (ti:(“Run”)) OR (ab:(“Jogging”)) OR (ti:(“Jogging”)) OR (ab:(“Walking”)) OR (ti:(“Walking”)) OR (ab:(“Cycling”)) OR (ti:(“Cycling”)) OR (ab:(“Swimming”)) OR (ti:(“Swimming”)) OR (((ab:(“Aerobic”)) OR (ti:(“Aerobic”)) OR (ab:(“Endurance”)) OR (ti:(“Endurance”)) OR (ab:(“Cardiovascular”)) OR (ti:(“Cardiovascular”)) OR (ab:(“Cardiopulmonar”)) OR (ti:(“Cardiopulmonar”)) OR (ab:(“Cardiorespiratory”)) OR (ti:(“Cardiorespiratory”)) OR (ab:(“Metabolic”)) OR (ti:(“Metabolic”)) OR (ab:(“Circuit”)) OR (ti:(“Circuit”)) OR (ab:(“Interval”)) OR (ti:(“Interval"))) AND ((ab:(“Exercise”)) OR (ti:(“Exercise”)) OR (ab:(“Training”)) OR (ti:(“Training”)) OR (ab:(“Workout”)) OR (ti:(“Workout)) OR (ab:(“Practice”)) OR (ti:(“Practice”)) OR (ab:(“Activity”)) OR (ti:(“Activity”)) OR (ab:(“Conditioning”)) OR (ti:(“Conditioning"))))) AND (ab:(“Trial”)) OR (ti:(“Trial”)) | 26^th^ Dec, 2023 | 3 |
| Google Scholar | - | Nº1 | (("Gonarthrosis" OR ("Knee" + ("Osteoarthritis" OR "Arthrosis" OR "Osteoarthrosis"))) + (“Running” OR “Swimming” OR “Walking” OR “Running” OR “Run” OR “Jogging” OR “Walking” OR “Cycling” OR “Swimming” OR “Aerobic” OR “Endurance” OR “Cardiovascular” OR “Cardiopulmonar” OR “Cardiorespiratory” OR “Metabolic” OR “Circuit”) + ("Exercise" OR "Training" OR “Workout” OR “Practice” OR “Activity” OR “Conditioning”) + “Trial”) -intitle:"review" -intitle:"systematic review" -intitle:"meta analysis" | 26^th^ Dec, 2023 | 980 |

Supplementary Material 2. Methods of Estimating Intensity of Cardiorespiratory Exercise by the ACSM 11^th^ Edition.

| Relative intensity | | | | Intensity (%O_2_max)  Relative to maximal exercise capacity in met | | | | Abs. Int. | Abs. Int. (met) by age | | |
| --- | --- | --- | --- | --- | --- | --- | --- | --- | --- | --- | --- |
| Intensity | %HRR or %VO_2_R | %HR max | %VO_2_ max | Perceived Exertion (Rating on 6-20 RPE Scale) | 20 METs %VO_2_ max | 10 METs %VO_2_ max | 5 METs %VO_2_ max | (METs) | Young (20-39 yr) | Middle Age (40-64 yr) | Older (≥65 yr) |
| Very light | <30 | <57 | <37 | Very light (RPE <9) | <34 | <37 | <44 | <2.0 | <2.4 | <2.0 | <1.6 |
| Light | 30-39 | 57-63 | 37-45 | Very light to fairly light (RPE 9-11) | 34-42 | 37-45 | 44-51 | 2.0-2.9 | 2.4-4.7 | 3.0-5.9 | 1.6-3.1 |
| Moderate | 40-59 | 64-76 | 46-63 | Fairly light to somewhat hard (RPE 12-13) | 43-61 | 46-63 | 52-67 | 3.0-5.9 | 4.8-7.1 | 5.9-9.3 | 4.7 |
| Vigorous | 60-89 | 77-95 | 64-90 | Somewhat hard to very hard (RPE 14-17) | 62-90 | 64-90 | 68-91 | 6.0-8.7 | 7.2-10.1 | 8.0-10.4 | 6.7 |
| Near-maximal to maximal | ≥90 | ≥96 | ≥91 | Very hard (RPE ≥18) | ≥91 | ≥91 | ≥92 | ≥8.8 | ≥10.2 | ≥8.5 | ≥6.8 |

Supplementary Material 3. Computation of Hedges’ g and its exact variance.

The standardized mean difference was computed using Hedges’ unbiased estimator $g_{t,ij}$, which adjusts Cohen’s $d_{t,ij}$ for small sample bias. First, Cohen’s d was calculated as the difference between experimental and control group means divided by the pooled standard deviation (Equation 3 in Hedges, 1982):

$$d_{t,ij}=\frac{x_{t,i}-x_{t,j}}{\sigma_{t,ij}^{*}}$$

With the pooled standard deviation defined as:

$$\sigma_{t,ij}^{*}=\sqrt{\frac{\left( N_{t,i}-1 \right){\cdot\sigma}_{t,ij}^{2}+\left( N_{t,j}-1 \right)\cdot\sigma_{t,j}^{2}}{N_{t,i}+N_{t,j}-2}}$$

To correct for bias in small samples, $d_{t,ij}$ was multiplied by the correction factor ${c(m)}_{t,ij}$, where $m_{t,ij}=N_{t,i}+ N_{t,j}-2$. Following the in Equation 5:

$${c(m)}_{t,ij}\approx1-\frac{3}{4m_{t,ij}-1}$$

This yielded the Hedges’ g $\left( g_{t,ij} \right)$ corrected estimator:

$$\Delta_{t,ij}={c(m)}_{t,ij}\cdot d_{t,ij}$$

The exact variance of $g_{t,ij}$ was estimated using the formula provided in Equation 6 of Hedges (1982):

$$\mathrm{Var}\left( g_{t,ij} \right)=\frac{\left[ {c\left( m \right)}_{t,ij} \right]^{2}\cdot m_{t,ij}\cdot\left( 1+ñ_{t,ij}\cdot g_{t,ij}^{2} \right)}{\left( N_{t,i}+N_{t,j}-4 \right)\cdotñ_{t,ij}}-g_{t,ij}^{2}$$

with $ñ_{t,ij}$ defined as:

$$ñ_{t,ij}=\frac{N_{t,i} \cdot N_{t,j}}{N_{t,i} + N_{t,j}}$$

Supplementary Material 4. Python syntax for computing Bayesian NMA, pairwise comparisons and funnel plots.

import numpy as np

import pandas as pd

import matplotlib.pyplot as plt

from matplotlib import cm, colormaps

import os

import pickle

import warnings

from tqdm import tqdm

import pickle

import time

from multiprocessing import cpu_count

import networkx as nx

import pymc3 as pm

import arviz as az

\# Auxiliary function to compute Hedge's G

\# the inputs are the efficacies, the errors, and the samples sizes respectively

def hedgesG(eff1, eff2, sd1, sd2, N1, N2):

m = N1 + N2 - 2

pool_sd = np.sqrt(((N1 - 1) * sd1**2 + (N2 - 1) * sd2**2) / m)

c = 1 - (3 / (4 * m - 1))

g12 = c * (eff2 - eff1) / pool_sd

err12 = np.sqrt(((N1 + N2) / (N1 * N2)) + (0.5 * g12**2 / (N1 + N2)))

return g12, err12

\# Main class of the module, actually running the Bayesian NMA

class koaBayesianNMA():

\# init function

def __init__(self, edgelist, edgelist_map=None):

\# the edgelist format is the same suggested by its map:

\# [study, treatment A, treatment B, trial-specific relative efficacy, its error,

\# weekly frequency of A, weekly frequency of B, n of weeks of A, n of weeks of B]

\# The covariates (freq and weeks) are set to zero in treatments where they are absent

if edgelist_map == None:

self.edgelist_map = {'study': 'study', 'T_a': 'T_a', 'T_b': 'T_b',

'delta_ab': 'delta_ab', 'std_ab': 'std_ab',

'freq_a': 'freq_a', 'freq_b': 'freq_b',

'weeks_a': 'weeks_a', 'weeks_b': 'weeks_b'}

elif any(('study' not in edgelist_map.keys(), 'T_a' not in edgelist_map.keys(),

'T_b' not in edgelist_map.keys(), 'delta_ab' not in edgelist_map.keys(),

'std_ab' not in edgelist_map.keys(), 'freq_a' not in edgelist_map.keys(),

'freq_b' not in edgelist_map.keys(), 'weeks_a' not in edgelist_map.keys(),

'weeks_b' not in edgelist_map.keys())):

raise Exception('Ill-defined edgelist_map!')

else:

self.edgelist_map = edgelist_map

self.edgelist = edgelist

self.treatments = np.unique(np.concatenate((edgelist[self.edgelist_map['T_a']],

edgelist[self.edgelist_map['T_b']])))

self.baseline = self.treatments[0]

self.n_d = self.treatments.shape[0] - 1

self.d_to_idx = dict(zip(self.treatments[1:], range(self.n_d)))

self.idx_to_d = dict(zip(range(self.n_d), self.treatments[1:]))

self.n_trials = self.edgelist.shape[0]

self.n_studies = np.unique(self.edgelist[self.edgelist_map['study']]).shape[0]

self.delta_shapes = []

self.X_matlist = []

self.delta_matlist = []

self.std_matlist = []

self.f_matlist, self.w_matlist = [], []

for study, ssdf in edgelist.groupby(self.edgelist_map['study']):

if ssdf.shape[0] > 1:

nodes = np.unique(np.concatenate((ssdf[self.edgelist_map['T_a']],

ssdf[self.edgelist_map['T_b']])))

baseline = nodes[0]

else:

baseline = None

deltas, stds, sub_X, sub_f, sub_w = [], [], [], [], []

for _, line in ssdf.iterrows():

fline, wline = np.zeros(self.n_d), np.zeros(self.n_d)

if baseline is None:

baseline = np.unique((line[self.edgelist_map['T_a']],

line[self.edgelist_map['T_b']]))[0]

xline = np.zeros(self.n_d)

if baseline != self.baseline:

xline[self.d_to_idx[baseline]] = -1

if line[self.edgelist_map['T_a']] != baseline:

xline[self.d_to_idx[line[self.edgelist_map['T_a']]]] = 1

f = line[self.edgelist_map['freq_a']]

fline[self.d_to_idx[line[self.edgelist_map['T_a']]]] = f

w = line[self.edgelist_map['weeks_a']]

wline[self.d_to_idx[line[self.edgelist_map['T_a']]]] = w

deltas.append(-line[self.edgelist_map['delta_ab']])

else:

xline[self.d_to_idx[line[self.edgelist_map['T_b']]]] = 1

f = line[self.edgelist_map['freq_b']]

fline[self.d_to_idx[line[self.edgelist_map['T_b']]]] = f

w = line[self.edgelist_map['weeks_b']]

wline[self.d_to_idx[line[self.edgelist_map['T_b']]]] = w

deltas.append(line[self.edgelist_map['delta_ab']])

sub_X.append(xline)

stds.append(line[self.edgelist_map['std_ab']])

self.delta_shapes.append(len(deltas))

self.delta_matlist.append(np.array(deltas))

self.std_matlist.append(np.array(stds))

self.X_matlist.append(np.array(sub_X))

fline = np.nan_to_num(fline)

wline = np.nan_to_num(wline)

self.f_matlist.append(fline)

self.w_matlist.append(wline)

self.model = None

\# builds the NMA model

def modelBuilder(self, noninfo_sigma=1e4, tau_max=5, simple=False, student_t=False):

\# noninfo_sigma is the sigma of the prior

\# tau_max is the upper limit of the tau flat prior [0, tau_max]

\# simple=True tells the function to include the covariates in the inference

\# student-t can be used to replace the Gaussians

self.model = pm.Model()

with self.model:

d_0 = pm.Normal("d_0", mu=np.zeros(self.n_d), shape=self.n_d, sigma=noninfo_sigma)

if simple:

c_f = np.zeros(self.n_d)

c_w = np.zeros(self.n_d)

else:

c_f = np.ones(self.n_d) * pm.Normal("c_f", mu=0, sigma=noninfo_sigma)

c_w = np.ones(self.n_d) * pm.Normal("c_w", mu=0, sigma=noninfo_sigma)

tau = pm.Uniform("tau", lower=0, upper=tau_max)

if student_t:

nu = pm.Uniform(f"nu", lower=0, upper=50)

for i in tqdm(range(len(self.X_matlist)), colour='green'):

xish = self.X_matlist[i].shape[0]

d = d_0 + pm.math.dot(c_w,self.w_matlist[i]) + pm.math.dot(c_f,self.f_matlist[i])

if xish == 1:

if student_t:

deltai = pm.StudentT(f"deltas_{i}", sigma=tau + self.std_matlist[i][0],

nu=nu, mu=pm.math.dot(self.X_matlist[i][0], d),

observed=self.delta_matlist[i][0])

else:

deltai = pm.Normal(f"deltas_{i}", sigma=tau + self.std_matlist[i][0],

mu=pm.math.dot(self.X_matlist[i][0], d),

observed=self.delta_matlist[i][0])

else:

sigmat = (np.ones((xish, xish)) + np.identity(xish)) * 0.5 * (tau ** 2)

varmat = np.diag(self.std_matlist[i]**2)

if student_t:

deltai = pm.MvStudentT(f"deltas_{i}", Sigma=sigmat+varmat, shape=xish,

nu=nu, observed=self.delta_matlist[i],

mu=pm.math.matrix_dot(self.X_matlist[i], d))

else:

deltai = pm.MvNormal(f"deltas_{i}", cov=sigmat+varmat, shape=xish,

mu=pm.math.matrix_dot(self.X_matlist[i], d),

observed=self.delta_matlist[i])

\# actually runs the model

def runBayes(self, niters=2500, burn_in=1000, chains=4, save_path=None, simple=False,

noninfo_sigma=1e4, tau_max=5, student_t=False, cores=None):

\# niters is the number of MCMC samples

\# burn_in is the number of tuning samples

\# chains is the number of MC chains

\# if save_path is not None, the trace is saved there

\# cores is the number of cores used by the model

\# all the other variables are the same as modelBuilder

startime = time.time()

self.modelBuilder(noninfo_sigma=noninfo_sigma, tau_max=tau_max,

simple=simple, student_t=student_t)

print('Starting to sample (this will take a while, especially after the prog-bar)...')

with self.model:

trace = pm.sample(niters, tune=burn_in, cores=cores,

chains=chains, return_inferencedata=True)

print('Done!')

elapstime = (time.time() - startime) // 60

print(f"\nBayesian sampling is over! [tte = {elapstime} mins]")

if save_path is not None:

pickle.dump(trace, open(save_path, 'wb'))

return trace

\# extracts the result table from the trace

def resulTable(trace, d_names, save_path=None, show=True, simple=False, student_t=False):

\# trace output from the main class

\# d_names are the names of the treatments, in the same order as they were in the main class

\# if save_path is not None, the trace is saved there

\# if show=True it prints the table

\# simple and student-t should match the ones in the main class

var_names = ['d_0', 'tau']

if student_t:

var_names.append('nu')

if not simple:

var_names.extend(['c_f', 'c_w'])

df = az.summary(trace, var_names=var_names, hdi_prob=0.95)

namecol = pd.Series(np.concatenate((d_names, var_names[1:])), name='variable')

df = pd.concat((namecol, df.reset_index(drop=True)), axis=1)

if save_path is not None:

df.to_csv(save_path, index=None)

if show:

display(df)

return df

\# generates the forest plot

def forestPlotter(df, n_d, title='', d_color='xkcd:scarlet', save_path=None,

xlim=5, fsize=25, good_effect=True):

\# df is the result table from the previous function

\# n_d is the number of treatments

\# title will be printed above the plot in the figure

\# d_color is the color of the treatmetn bars

\# if save_path is not None, the trace is saved there

\# xlim is the limit of the x-axis (treatment efficacy)

\# fsize is the fontsize

\# if good_effect=True a positive value of d favors intervention

\# if good_effect=False a positive value of d favors "No Treatment"

figa, ax = plt.subplots(1, 1, figsize=(16, n_d + 1))

hdi = 0.5 * (df['hdi_97.5%'].iloc[:n_d] - df['hdi_2.5%'].iloc[:n_d])

dy = [*reversed(range(df.shape[0] - n_d, df.shape[0]))]

xtreats = df['mean'].iloc[:n_d]

idx = np.argsort(xtreats)

ax.errorbar(xtreats[idx], dy, xerr=hdi[idx], color=d_color, linewidth=2,

zorder=5, linestyle='')

ax.scatter(xtreats[idx], dy, marker='s', s=150, facecolor='white',

edgecolor=d_color, zorder=50, linewidth=2)

renames = {'EP': 'ED', 'LMV': 'AE_LIGHT_MOD_VIG', 'LM': 'AE_LIGHT_MOD', 'M': 'AE_MOD',

'MV': 'AE_MOD_VIG', 'NT': 'NT', 'RE': 'RE', 'V': 'AE_VIG',

'REBLM': 'RE_BEHAV', 'REASE': 'RE_AE_STRETCH'}

for i, di in zip(idx, dy):

ax.text(-xlim*1.67, di, renames[df['variable'].iloc[i]],

ha='left', va='center', fontsize=fsize)

ax.text(xlim*1.2, di, np.round(df['mean'].iloc[i], 2),

ha='right', va='center', fontsize=fsize)

ax.text(xlim*1.67, di, f"[{df['hdi_2.5%'].iloc[i]:.2f}; {df['hdi_97.5%'].iloc[i]:.2f}]",

ha='right', va='center', fontsize=fsize)

ax.text(-xlim*1.67, dy[0]+1, 'Treatments', fontweight='bold',

ha='left', va='center', fontsize=fsize)

ax.text(0, dy[0]+1, title, fontweight='bold',

ha='center', va='center', fontsize=fsize)

ax.text(xlim*1.2, dy[0]+1, 'SMD', fontweight='bold',

ha='right', va='center', fontsize=fsize)

ax.text(xlim*1.67, dy[0]+1, '95%-CI', fontweight='bold',

ha='right', va='center', fontsize=fsize)

if good_effect:

ax.text(-xlim, dy[-1]-1.5, 'Favors No Treatment', fontsize=fsize, ha='left', va='center')

ax.text(xlim, dy[-1]-1.5, 'Favors Intervention', fontsize=fsize, ha='right', va='center')

else:

ax.text(-xlim, dy[-1]-1.5, 'Favors Intervention', fontsize=fsize, ha='left', va='center')

ax.text(xlim, dy[-1]-1.5, 'Favors No Treatment', fontsize=fsize, ha='right', va='center')

ax.axvline(0, color='black', linewidth=1.5, zorder=10, linestyle='-.')

ax.set_yticks(dy)

ax.set_yticklabels('', fontsize=17)

ax.tick_params(axis='x', labelsize=fsize)

ax.set_ylim(dy[-1] - 0.5, dy[0] + 0.5)

ax.set_xlim(-xlim, xlim)

ax.set_facecolor('white')

ax.yaxis.set_visible(False)

ax.grid(False)

for spine in ['top', 'left', 'right']:

ax.spines[spine].set_visible(False)

if save_path is not None:

plt.savefig(save_path, bbox_inches='tight')

plt.show()

\# infers direct and indirect estimates

def directIndirectEstimator(edgelist, resultable, niters=10000, burn_in=20000, simple=True,

save_path=None):

\# edgelist should be in the same format as the main class

\# resultable is the output of the same-name function

\# niters is the number of MCMC samples

\# burn_in is the number of tuning samples

\# if save_path is not None, the trace is saved there

nodelabels = {'Education protocol': 'EP', 'Light-to-Moderate-to-Vigorous': 'LMV',

'Light-to-Moderate': 'LM', 'Moderate': 'M', 'Moderate-to-Vigorous': 'MV',

'0_ No treatment': 'NT', 'Resistance exercise': 'RE', 'Vigorous': 'V',

'Resistance exercise and Behavioral-lifestyle modification': 'REBLM',

'Resistance, Aerobic, and Stretching exercises': 'REASE'}

nodes = np.unique(np.concatenate((edgelist['T_a'], edgelist['T_b'])))

N, M = len(nodes), edgelist.shape[0]

d_nma = np.concatenate(([0], resultable['mean'].values[:N-1]))

std_nma = np.concatenate(([0], resultable['sd'].values[:N-1]))

\# d5_nma = np.concatenate(([0], resultable['hdi_2.5%'].values[:N-1]))

\# d95_nma = np.concatenate(([0], resultable['hdi_97.5%'].values[:N-1]))

if simple:

tau_nma = resultable['mean'].values[-1]

tausd_nma = resultable['mean'].values[-1]

else:

tau_nma = resultable['mean'].values[-3]

tausd_nma = resultable['mean'].values[-3]

G0 = nx.MultiGraph()

G0.add_nodes_from(nodes)

print(f"Nodes: {[nodelabels[n] for n in nodes]}\n")

final_df = []

final_cols = ['node_a', 'node_b', 'direct_est', 'direct_std', 'direct_2.5%',

'direct_97.5%', 'indirect_est', 'indirect_std', 'indirect_2.5%',

'indirect_97.5%', 'indirect_tau', 'indirect_tau_std', 'mixed_est',

'mixed_std', 'mixed_2.5%', 'mixed_97.5%', 'mixed_tau', 'mixed_tau_std']

totals = N * (N - 1) // 2

c = 1

start = time.time()

for i in range(N):

for j in range(i + 1, N):

print(f"EVALUATING {nodelabels[nodes[i]]}-{nodelabels[nodes[j]]} ({c} / {totals})")

G = G0.copy()

ind_df = []

dir_df = []

for _, line in edgelist.iterrows():

if line['T_a'] == nodes[i] and line['T_b'] == nodes[j]:

dir_df.append(line.values)

elif line['T_a'] == nodes[j] and line['T_b'] == nodes[i]:

dir_df.append(line.values)

else:

G.add_edge(line['T_a'], line['T_b'])

ind_df.append(line.values)

if len(ind_df) == M:

dir_d = float('nan')

dir_std = float('nan')

dir_5 = float('nan')

dir_95 = float('nan')

ind_d = d_nma[j] - d_nma[i]

ind_std = np.sqrt(std_nma[j]**2 + std_nma[i]**2)

ind_5 = ind_d - 1.96 * ind_std

ind_95 = ind_d + 1.96 * ind_std

ind_tau = tau_nma

ind_tausd = tausd_nma

elif not nx.is_connected(G):

dir_df = pd.DataFrame(dir_df, columns=edgelist.columns)

dir_d = np.average(dir_df['delta_ab'], weights=1/dir_df['std_ab'])

dir_std = (np.sum(dir_df['std_ab']**-1))**-1

dir_5 = dir_d - 1.96 * dir_std

dir_95 = dir_d + 1.96 * dir_std

ind_d = float('nan')

ind_std = float('nan')

ind_5 = float('nan')

ind_95 = float('nan')

ind_tau = float('nan')

ind_tausd = float('nan')

else:

dir_df = pd.DataFrame(dir_df, columns=edgelist.columns)

dir_d = np.average(dir_df['delta_ab'], weights=1/dir_df['std_ab'])

dir_std = (np.sum(dir_df['std_ab']**-1))**-1

dir_5 = dir_d - 1.96 * dir_std

dir_95 = dir_d + 1.96 * dir_std

ind_df = pd.DataFrame(ind_df, columns=edgelist.columns)

ind_nma = UnivariateBayesianNMA(pain_df)

trace = ind_nma.runBayes(niters=niters, burn_in=burn_in, chains=4, simple=simple,

student_t=False, noninfo_sigma=10, save_path=None)

ind_table = resulTable(trace, nodes, show=False, simple=simple,

student_t=False, save_path=None)

d_vec = np.concatenate(([0], ind_table['mean'].values[:N-1]))

std_vec = np.concatenate(([0], ind_table['sd'].values[:N-1]))

ind_d = d_vec[j] - d_vec[i]

ind_std = np.sqrt(std_vec[j]**2 + std_vec[i]**2)

ind_5 = ind_d - 1.96 * ind_std

ind_95 = ind_d + 1.96 * ind_std

if simple:

ind_tau = ind_table['mean'].values[-1]

ind_tausd = ind_table['mean'].values[-1]

else:

ind_tau = ind_table['mean'].values[-3]

ind_tausd = ind_table['mean'].values[-3]

nma_d = d_nma[j] - d_nma[i]

nma_std = np.sqrt(std_nma[j]**2 + std_nma[i]**2)

nma_5 = nma_d - 1.96 * nma_std

nma_95 = nma_d + 1.96 * nma_std

final_df.append([nodelabels[nodes[i]], nodelabels[nodes[j]], dir_d, dir_std, dir_5,

dir_95, ind_d, ind_std, ind_5, ind_95, ind_tau, ind_tausd,

nma_d, nma_std, nma_5, nma_95, tau_nma, tausd_nma])

c += 1

print()

final_df = pd.DataFrame(final_df, columns=final_cols)

if save_path is not None:

final_df.to_csv(save_path)

display(final_df.head(10))

print(f"DIRECT-INDIRECT ESTIMATION DONE [tte = {(time.time() - start) // 60} mins]")

def dirIndZ(table_path):

\# this function performs the Z-test between direct and indirect estimates

\# adding the result as a column to the table generated by the function above

\# the table path itself is the input

df = pd.read_csv(table_path)

sigma = np.sqrt(df['direct_std'].values**2 + df['indirect_std'].values**2)

z = (df['direct_est'].values - df['indirect_est'].values) / sigma

df['Z_dir_ind'] = 2 * norm.cdf(-np.abs(z))

df.to_csv(table_path, index=None)

\# generates the funnel plots

def funnelPlots(pain=True, simple=True, ylims=(None, None), xlims=(None, None)):

\# if pain is true it imports the pain networks, else the walking-func ones

\# if simple it imports the table with no covariates, else with covariates

\# ylims and xlims delimit the axes in the plot

studict = {'Beckwee 2015': 0.666, 'de Almeida 2019': 0.1, 'de Almeida 2020': 0.2,

'Ettinger 1997': 0.3, 'Lim 2010': 0.4, 'Keogh 2018': 0.5, 'Mangione 1997': 0.6,

'Moghadam 2017': 0.75, 'Oistad 2023': 0.8, 'Salacinski 2012': 0.9,

'Samut 2015': 0.99}

nodelabels = {'Education protocol': 'EP', 'Light-to-Moderate-to-Vigorous': 'LMV',

'Light-to-Moderate': 'LM', 'Moderate': 'M', 'Moderate-to-Vigorous': 'MV',

'No treatment': 'NT', 'Resistance exercise': 'RE', 'Vigorous': 'V',

'Resistance exercise and Behavioral-lifestyle modification': 'REBLM',

'Resistance, Aerobic, and Stretching exercises': 'REASE',

'0_ No treatment': 'NT'}

aerobics = ['LMV', 'LM', 'M', 'MV', 'V']

if pain:

vartag = 'pain'

indict = {0: 'Beckwee 2015', 1: 'de Almeida 2019', 2: 'Ettinger 1997', 3: 'Lim 2010',

4: 'Oistad 2023', 5: 'Salacinski 2012', 6: 'Samut 2015'}

else:

vartag = 'walk'

indict = {0: 'de Almeida 2020', 1: 'Ettinger 1997', 2: 'Keogh 2018', 3: 'Mangione 1997',

4: 'Moghadam 2017', 5: 'Salacinski 2012', 6: 'Samut 2015'}

if simple:

baytag = 'simple'

else:

baytag = 'covars'

raw_df = pd.read_csv(f"{out_path}{vartag}_edgelist.csv")

nma_df = pd.read_csv(f"{out_path}{vartag}_1v-{baytag}-gauss_resultable.csv")

figa, ax = plt.subplots(1, 1, figsize=(9, 6))

ax.yaxis.set_inverted(True)

olim = 2.5

ax.plot([0, olim], [0, 1.96 * olim], color='black', linestyle='--', alpha=0.5)

ax.plot([0, -olim], [0, 1.96 * olim], color='black', linestyle='--', alpha=0.5)

ax.plot([0, 0], [0, 1.96 * olim], color='black', linestyle='--', alpha=0.5)

for study, sdf in raw_df.groupby('study'):

ax.scatter([], [], color=cm.gist_ncar_r(studict[indict[study]]), zorder=25,

label=indict[study], edgecolor='black', linewidth=0.5)

for _, line in sdf.iterrows():

if line['T_a'] == '0_ No treatment':

da = 0

stda = 0

else:

da = nma_df[nma_df['variable'] == nodelabels[line['T_a']]]['mean'].values[0]

stda = nma_df[nma_df['variable'] == nodelabels[line['T_a']]]['sd'].values[0]

if line['T_b'] == '0_ No treatment':

db = 0

stdb = 0

else:

db = nma_df[nma_df['variable'] == nodelabels[line['T_b']]]['mean'].values[0]

stdb = nma_df[nma_df['variable'] == nodelabels[line['T_b']]]['sd'].values[0]

x = db - da - line['delta_ab']

y = np.sqrt(line['std_ab']**2 + stda**2 + stdb**2)

if nodelabels[line['T_a']] in aerobics and nodelabels[line['T_b']] in aerobics:

symbol = '^'

elif nodelabels[line['T_a']] not in aerobics and nodelabels[line['T_b']] not in aerobics:

symbol = 'o'

else:

symbol = 's'

ax.scatter(x, y, color=cm.gist_ncar_r(studict[indict[study]]), zorder=25,

marker=symbol, edgecolor='black', linewidth=0.5)

if y < np.abs(x * 1.96):

print(indict[study], nodelabels[line['T_a']], nodelabels[line['T_b']],

np.round(db - da, 3), np.round(line['delta_ab'], 3), np.round(y, 3))

ax.set_xlim(*xlims)

ax.set_ylim(*ylims)

ax.set_xlabel("Difference between network estimate and study-specific effect")

ax.set_ylabel("Error associated to the difference on the x-axis")

ax.legend(ncol=4, loc='upper center')

plt.savefig(f"{out_path}funnel-plots_{vartag}_{baytag}.pdf", bbox_inches='tight')

plt.show()
