## Supplementary material for "How intense is effective? Exploring aerobic exercise intensity for knee osteoarthritis through a Bayesian network meta-analysis": T1 Summary

Table 1. Summary information from the included studies

| Study | Population | Groups | Prescription parameters and monitorization | Outcome measures and instruments | Results (post-immediate) | | | |
| --- | --- | --- | --- | --- | --- | --- | --- | --- |
|  |  |  |  |  | **Narrative** | | | **Effect direction** |
| Arrieiro et al., 2019  *Parallel RCT* | **KOA diagnostic criteria:** Clinical and radiological according to ACR *(only radiological reference)*  **Presence of pain:** Probable but unclear | **Vigorous intensity AE** (n_allo_=8, n_an_=8):  Age: 68.6 ± 6; F (n): 8; BMI: 28.7 ± 5.39; Diabetes: n/a; Hypertension: n/a  **Vigorous intensity AE** (n_allo_=8, n_an_=8):  Age: 67 ± 3; F(n): 8; BMI: 27.56 ± 2.05; Diabetes: n/a; Hypertension: n/a | **Continuous Land-based walking:**  **Condit. Intensity:** Moderate (70-75% HR_max_)  **Final intensity:** Vigorous (75-80%, 75-80% and 80-85% HR_max_)  **Monit. Device:** Polar, model F4  **Volume:** Total duration (45, 50, 55, 60 and 65 min); Stimuli duration (35, 40, 45, 50 and 55 min); 3 sess/week for 12 weeks  **Continuous Walter-based walking:**  **Condit. Intensity:** Moderate (70-75% HR_max_)  **Final intensity:** Vigorous (75-80%, 75-80% and 80-85% HR_max_)  **Monit. Device:** Polar, model F4  **Volume:** Total duration (45, 50, 55, 60 and 65 min); Stimuli duration (35, 40, 45, 50 and 55 min); 3 sess/week for 12 weeks | Pain intensity (WOMAC pain subscale) | No relevant differences between groups were found | | | Vig AE Water-based ≈ Vig AE Land-based |
|  |  |  |  | Walking performance (6MWT) | No relevant differences between groups were found | | | Vig AE Water-based ≈ Vig AE Land-based |
|  |  |  |  | Stiffness (WOMAC- Stiffness) | No relevant differences between groups were found | | | Vig AE Water-based ≈ Vig AE Land-based |
| Bavardi Moghadam & Shojaedin 2017  *Parallel RCT* | **KOA diagnostic criteria:** Clinical and radiological criteria *(no followed guidelines)*  **Presence of pain:** Yes, pain during the previous 6 months | **Moderate-to-Vigorous AE** (n_allo_=10, n_an_=9):  Age: 64.66 ± 3.46; F (n=10); BMI: 24.93 ±0.92; Diabetes n/a; Hypertension: n/a  **No treatment** (n_allo_=10, n_an_=9):  Age: 68.77 ± 2.99; F (n=10); BMI: 25.39 ± 1.87; Diabetes n/a; Hypertension: n/a | **Continuous walking:**  **Condit. Intensity:** 60%. Light  **Final intensity:** Moderate-to-Vigorous; 65-75% HR_max_  **Monit. Device:** Pulsometer (No model info.)  **Volume:** Total duration (No info.); Stimuli duration (20-30min); 3 sess/week for 8 weeks  **No treatment:**  **Volume:** 8 weeks | Walking performance (6MWT) | There was a significant improvement in the walking group compared to the control group | | | Mod-Vig AE > No Treatment |
|  |  |  |  | Function in sit to stand and linear walk (chair-to-stand and walk 15.2m) | There was a significant improvement in the walking group compared to the control group | | | Mod-Vig AE < No Treatment |
| Beckwee et al., 2015  *Parallel RCT* | **KOA diagnostic criteria:** Clinical and radiological according to ACR (Altman, 1995)  **Presence of pain:** Yes, knee pain in the previous 30 days | **Vigorous AE** (n_allo_=19, n_an_=17):  Age (mdn[IQR]): 60[10]; F (n): 8; BMI (mdn[IQR]): 27.9[5.2]; Diabetes: n/a; Hypertension (mdn[IQR] n): none  **Resistance exercise** (n_allo_=19, n_an_=15):  Age (mdn[IQR]): 61[10]; F (n): 13; BMI (mdn[IQR]): 27.1[7.9]; Diabetes: n/a; Hypertension (mdn[IQR] n): 1[5] | **Continuous walking:**  **Condit. Intensity:** No  **Final intensity:** Vigorous (14-17 Borg)  **Monit. Device:** Borg scale (6-20 points)  **Volume:** Total duration (No info.); Stimuli duration (40 min); 3 sess/week for 18 weeks  **Resistance exercise:**  **Volume:** Total duration (45 min); Stimuli duration (No info.); 3 sess/week; 18 weeks | Pain intensity during exercise (NRS post-pre) | | No relevant differences between groups were found | Vig AE ≈ Resistance exercise | |
|  |  |  |  | Maximum pain intensity perceived (NRS-max24) | | No relevant differences between groups were found | Vig AE ≈ Resistance exercise | |
| Casilda-López et al., 2017  *Parallel RCT* | **KOA diagnostic criteria:** Clinical *(no guidelines followed)*  **Presence of pain:** Probable but unclear | **Vigorous intensity AE** (n_allo_=17, n_an_=17):  Age: 66 ± 6.35; F (n=17); BMI: 33.65 ± 3.04; Diabetes: n/a; Hypertension: n/a  **Vigorous intensity AE** (n_allo_=17, n_an_=17):  Age: 65.62 ± 7.15; F (n=17); BMI: 31.69 ± 2.44; Diabetes: n/a; Hypertension: n/a | **Continuous aquatic global exercises:**  **Condit. Intensity:** No  **Final intensity:** Vigorous (4-6 Borg CR-10)  **Monit. Device:** Borg scale CR-10  **Volume:** Total duration (45 min); Stimuli duration (21 min); 3 sess/week for 8 weeks  **Intervallic aquatic dance:**  **Condit. Intensity:** No  **Final intensity:** Vigorous (4-6 Borg CR-10)  **Monit. Device:** Borg scale CR-10  **Volume:** Total duration (45 min); Stimuli duration (21 min); 3 sess/week for 8 weeks | Pain (WOMAC- Pain) | Reduction in aquatic dance group compared to the global aquatic exercise group | | | Vig AE Aquatic dance < Vig AE Global exercise |
|  |  |  |  | Walking performance (6MWT (m)) | An increase was found in aquatic dance group compared to the global aquatic exercise group | | | Vig AE Aquatic dance > Vig AE Global exercise |
|  |  |  |  | Knee stiffness (WOMAC- Stiffness) | No differences between groups were found | | | Vig AE Aquatic dance ≈ Vig AE Global exercise |
|  |  |  |  | Disability associated with KOA (WOMAC total score) | Reduction in aquatic dance group compared to the global aquatic exercise group | | | Vig AE Aquatic dance < Vig AE Global exercise |
| De Almeida., 2019  *Parallel RCT* | **KOA diagnostic criteria:** Clinical and radiological according to ACR (Altman, 1986)  **Presence of pain:** Yes, knee pain the previous week of ≥ 4 points on VAS (0-10) | **Light-to-moderate-to-vigorous AE** (n_allo_=22, n_an_=20):  Age: 55.6 ± 5.3; F (n=15); BMI: 26 ± 3.08; Diabetes: n/a; Hypertension: n/a  **Resistance training** (n_allo_=22, n_an_=21):  Age: 55.2 ± 7.4; F (n=16); BMI: 26 ± 3.14; Diabetes: n/a; Hypertension: n/a  **Educational protocol** (n_allo_=22, n_an_=20):  Age: 53.8 ± 7.7; F (n=16); BMI: 27 ± 2.7; Diabetes: n/a; Hypertension: n/a | **Intervallic full-body circuit:**  **Condit. Intensity:** Very light (<54% HR_max_ / 6-10 Borg)  **Final intensity:** Light-to-moderate-to-vigorous (<54% HR_max_ / 6-10 Borg progressed to 55-69% HR_max_ / 11-14 Borg, progressed to >70% HR_max_ / 15-20 Borg)  **Monit. Device:** No info for HR, Borg scale (6-20 points)  **Volume:** Total duration (30-45 min); Stimuli duration (20-35 min); 3 sess/week for 14 weeks  **Resistance exercise:**  **Volume:** Total duration (60 min); Stimuli duration (50 min); 3 sess/week for 14 weeks  **Education protocol:**  **Volume:** Total duration (60 min); Stimuli duration (No info.); 0.57 sess/week (2 sess/month) for 14 weeks | Pain intensity (VAS) | Greater reduction in the circuit training group compared to the educational group  No differences between aerobic and resistance group | | | Light-Mod-Vig AE < Educational group  Light-Mod-Vig AE ≈ Resistance exercise |
| De Almeida et al., 2020  *Parallel RCT*  *Same sample as Almeida et al. 2019* | **KOA diagnostic criteria:** Clinical and radiological according to ACR (Altman, 1986)  **Presence of pain:** Yes, knee pain the previous week of ≥ 4 points on VAS (0-10) | **Light-to-moderate-to-vigorous AE** (n_allo_=22, n_an_=20):  Age: 55.6 ± 5.3; F (n=15); BMI: 26 ± 3.08; Diabetes: n/a; Hypertension: n/a  **Resistance training** (n_allo_=22, n_an_=21):  Age: 55.2 ± 7.4; F (n=16); BMI: 26 ± 3.14; Diabetes: n/a; Hypertension: n/a  **Educational protocol** (n_allo_=22, n_an_=20):  Age: 53.8 ± 7.7; F (n=16); BMI: 27 ± 2.7; Diabetes: n/a; Hypertension: n/a | **Intervallic full-body circuit:**  **Condit. Intensity:** Very light (<54% HR_max_ / 6-10 Borg).  **Final intensity:** Light-to-moderate-to-vigorous (<54% HR_max_ / 6-10 Borg progressed to 55-69% HR_max_ / 11-14 Borg, progressed to >70% HR_max_ / 15-20 Borg)  **Monit. Device:** No info for HR, Borg scale (6-20 points)  **Volume:** Total duration (30-45 min); Stimuli duration (20-35 min); 3 sess/week for 14 weeks  **Resistance exercise:**  **Volume:** Total duration (60 min); Stimuli duration (50 min); 3 sess/week for 14 weeks  **Education protocol:**  **Volume:** Total duration (60 min); Stimuli duration (No info.); 0.57 sess/week (2 sess/month) for 14 weeks | Pain intensity (WOMAC- Pain) | Greater reduction in the circuit training group compared to the educational group  No differences between aerobic and resistance group | | | Light-Mod-Vig AE < Educational group  Light-Mod-Vig AE ≈ Resistance exercise |
|  |  |  |  | Walking performance (40mWT (m/s)) | No differences between groups were found after treatment  No differences between aerobic and resistance group | | | Light-Mod-Vig AE ≈ Educational group  Light-Mod-Vig AE ≈ Resistance exercise |
|  |  |  |  | Function in sit to stand (30sSTS) | Greater increase in the circuit training group compared to the educational group  No differences between aerobic and resistance group | | | Light-Mod-Vig AE > Educational group  Light-Mod-Vig AE ≈ Resistance exercise |
|  |  |  |  | Stiffness (WOMAC- Stiffness) | Greater reduction in the circuit training group compared to the educational group  No differences between aerobic and resistance group | | | Light-Mod-Vig AE < Educational group  Light-Mod-Vig AE ≈ Resistance exercise |
|  |  |  |  | Disability associated with KOA (WOMAC- Total) | Greater reduction in the circuit training group compared to the educational group  No differences between aerobic and resistance group | | | Light-Mod-Vig AE < Educational group  Light-Mod-Vig AE ≈ Resistance exercise |
| Ettinger et al., 1997  *Parallel RCT* | **KOA diagnostic criteria:** Clinical and radiological *(no guidelines followed)*  **Presence of pain:** Yes**,** pain on most days and pain during activities | **Moderate-to-Vigorous AE** (n_allo_=144, n_an_=117):  Age: 69 ± 6; F (n=99); BMI: n/a; Obesity (n=72); Diabetes (n=10); Hypertension (n=58)  **Resistance exercise** (n_allo_=146, n_an_=120):  Age: 68 ± 6; F (n=107); BMI: n/a; Obesity (n=72); Diabetes (n=14); Hypertension (n=61)  **Education protocol** (n_allo_=149, n_an_=127):  Age: 69 ± 6; F (n=102); BMI: n/a; Obesity (n=87); Diabetes (n=16); Hypertension (n=75) | **Continuous walking:**  **Condit. Intensity:** No  **Final intensity:** Moderate-to-Vigorous (50-70% HRR)  **Monit. Device:** No info.  **Volume:** Total duration (60min); Stimuli duration (40 min); 3 sess/week for 79 weeks  **Resistance exercise:**  **Volume:** Total duration (60 min); Stimuli duration (40 min); 3 sess/week; 79 weeks  **Education protocol:**  **Volume:** Total duration (110 min); Stimuli duration (No info.); 0.25 sess/week (1 sess/month); 79 weeks | Pain intensity- during walking and transfers (1-6 scale) | Aerobic group showed less pain after intervention compared to education group  No post-intervention comparisons between aerobic and resistance groups reported | | | Mod-Vig AE < Education group  No info. |
|  |  |  |  | Walking performance (6MWT) | Aerobic group walked greater distance after intervention compared to education group  No post-intervention comparisons between aerobic and resistance groups reported | | | Mod-Vig AE > Education group  No info. |
| Keogh et al., 2018  *Parallel RCT* | **KOA diagnostic criteria:** No info concerning if clinical and/or radiological *(conducted by a surgeon, and no guidelines followed)*  **Presence of pain:** Unclear | **Vigorous AE** (n_allo_=15, n_an_=9):  Age: 59.1 ± 6.7; F (n=6); BMI: 27.0 ± 4; Diabetes n/a; Hypertension: n/a  **Moderate AE** (n_allo_=12, n_an_=8):  Age: 66.1 ± 8.8; F (n=7); BMI: 28.2 ±6.9; Diabetes: n/a; Hypertension: n/a | **Intervallic cycling:**  **Condit. Intensity:** No  **Final intensity:** Vigorous *(“an intensity in which you felt it was quite difficult to complete sentences during the exercise”)*  **Monit. Device:** Talk test.  **Volume:** Total duration (25 min); Stimuli duration (11.25 min, involving 5 intervals of 45 s high intensity bouts and 90 s recovery); 4 sess/week for 8 weeks  **Continuous cycling:**  **Condit. Intensity:** No  **Final intensity:** Moderate *(“an intensity in which you are able to speak in complete sentences during exercise”)*  **Monit. Device:** Talk test.  **Volume:** Total duration (25 min); Stimuli duration (20 min); 4 sess/week for 8 weeks | Walking performance (normal walk speed in 3.66 m) | No differences were found after treatment between groups | | | Vig AE ≈ Mod AE |
|  |  |  |  | Function in sit to stand (30s Chair-to-Stand) | No differences were found after treatment between groups | | | Vig AE ≈ Mod AE |
|  |  |  |  | Function in sit to stand and walking (TUG) | Significant reduction after treatment in the HIIT group compared to MICT | | | Vig AE < Mod AE |
|  |  |  |  | Disability associated with KOA (WOMAC- total) | No differences were found after treatment between groups | | | Vig AE ≈ Mod AE |
| Lim et al., 2010  *Parallel RCT* | **KOA diagnostic criteria:** Radiological according to KL criteria *(grades ≥2)*  **Presence of pain:** Probable but unclear | **Moderate-to-Vigorous AE** (n_allo_=26, n_an_=24):  Age: 65.7 ± 8.9; F (n=23); BMI: 27.9 ± 1.5; Diabetes n/a; Hypertension: n/a  **Resistance, Aerobic, and Stretching exercises** (n_allo_=25, n_an_=22):  Age: 67.7 ± 7.7; F (n=21); BMI: 27.6 ± 7.7; Diabetes n/a; Hypertension: n/a  **Resistance exercise and Behavioral-lifestyle modification** (n_allo_=24, n_an_=20):  Age: 63.3 ± 5.3; F (n=21); BMI: 27.7 ± 2; Diabetes n/a; Hypertension: n/a | **Continuous aquatic exercise:**  **Condit. Intensity:** No  **Final intensity:** Moderate-to-Vigorous (≥65% HR_max_)  **Monit. Device:** No info.  **Volume:** Total duration (40min); Stimuli duration (30 min); 3 sess/week for 8 weeks  **Resistance, Aerobic, and Stretching exercises:**  **Volume:** Total duration (40 min); Stimuli duration (30 min); 3sess/week for 8 weeks  **Resistance exercise and Behavioural-lifestyle modification:**  **Volume:** Total duration (No info); Stimuli duration (No info); No info for sess/week; 8 weeks | Pain intensity (BPI scale) | No differences between aerobic and the group combining resistance, aerobic and stretching exercises  No differences between aerobic and the group combining resistance, and behavioral modifications | | | Mod-Vig AE ≈ Resistance, aerobic and stretching  Mod-Vig AE ≈ Resistance exercise and behavioral |
|  |  |  |  | Disability associated with KOA (WOMAC- total) | Greater decrease of disability in the water based aerobic exercise compared to resistance and behavioural after treatment  No differences after treatment were found between aquatic aerobic exercise and resistance, aerobic and stretching | | | Mod-Vig AE < Resistance exercise and behavioral  Mod-Vig AE ≈ Resistance, aerobic and stretching |
| Mangione et al., 1999  *Parallel RCT* | **KOA diagnostic criteria:** Clinical and radiological according to ACR (Altman, 1986)  **Presence of pain:** Yes | **Vigorous AE** (n_allo_=19, n_an_*):  Age: 71.1 ± 7.7; F (n=14); BMI: 29.63 ± 5.18; Diabetes n/a; Hypertension: n/a  **Light-to-moderate AE** (n_allo_=20, n_an_*):  Age: 71.0 ± 6.2; F (n=12); BMI: 29.08 ± 5.07; Diabetes n/a; Hypertension: n/a | **Continuous stationary cycling:**  **Condit. Intensity:** No  **Final intensity:** Vigorous (70% HRR)  **Monit. Device:** Polar Electro, Inc., Port Washington, NY  **Volume:** Total duration (60min); Stimuli duration (25 min); 3 sess/week for 10 weeks  **Continuous Stationary cycling:**  **Condit. Intensity:** No  **Final intensity:** Light-to-Moderate (40% HRR)  **Monit. Device:** Polar Electro, Inc., Port Washington, NY  **Volume:** Total duration (60 min); Stimuli duration (25 min); 3 sess/week for 10 weeks | Walking performance (6MWT, normal walk speed in 3.87m and fast walk speed in 3.87 m) | No differences found between groups after intervention | | | Vig AE ≈ Light-mod AE |
|  |  |  |  | Function in sit to stand (10 Chair-to-stand) | No differences found between groups after intervention | | | Vig AE ≈ Light-mod |
| Messier et al., 1997  *Sub-sample of Ettinger 1997*  *Parallel RCT* | **KOA diagnostic criteria:** Clinical and radiological according to ACR (Altman, 1986)  **Presence of pain:**  Yes, pain on most days of the month | **Moderate-to-Vigorous AE** (n_allo_=33, n_an_=33):  Age: 70.3 ± 1.3; F (n=27); BMI: 31.4 ±1.0; Diabetes n/a; Hypertension: n/a  **Resistance exercise** (n_allo_=34, n_an_=34):  Age: 67.2 ± 0.9; F (n=23); BMI: 30.1 ± 0.9; Diabetes n/a; Hypertension: n/a  **Education protocol** (n_allo_=36, n_an_=36):  Age: 69.2 ± 1.0; F (n=28); BMI: 32.5 ± 0.9; Diabetes n/a; Hypertension: n/a | **Continuous walking:**  **Condit. Intensity:** No  **Final intensity:** Moderate-to-Vigorous (50-85% HRR)  **Monit. Device:** No info.  **Volume:** Total duration (50 min); Stimuli duration (40 min); 3 sess/week for 79 weeks^†^  **Resistance exercise:**  **Volume:** Total duration (No info.); Stimuli duration (No info.) 3sess/week; 79 weeks^†^  **Education protocol:**  **Volume:** Total duration (60 min); Stimuli duration (No info.); 0.5 phone call/week; 79 weeks^†^ | Pain intensity during ambulation and during transfers (1-6 scale) | No differences found between aerobic and education  No differences found between aerobic and resistance exercise | | | Mod-Vig AE ≈ Education  Mod-Vig AE ≈ Resistance |
|  |  |  |  | Walking performance (22mWT) (cm/s) | There was a significant increase in walking speed in the aerobic group compared to the education group  No differences found between aerobic and resistance group | | | Mod-Vig AE > Education  Mod-Vig AE ≈ Resistance |
| Øiestad et al., 2023  *Parallel RCT* | **KOA diagnostic criteria:** Clinical and radiological according to ACR (Altman, 1986; Kellgren, 1957)  **Presence of pain:**  Yes, pain on most days of the month | **Moderate-to-Vigorous AE** (n_allo_=55, n_an_=42):  Age: 57.3 ± 7.1; F (n=28); BMI: 29.4 ± 4.4; Diabetes n/a; Hypertension: n/a  **Resistance exercise** (n_allo_=57, n_an_=49):  Age: 57.6 ± 6.6; F (n=30); BMI: 28.9 ± 4.3; Diabetes n/a; Hypertension: n/a  **No treatment** (n_allo_=56, n_an_=45):  Age: 57.8 ± 7.4; F (n=24); BMI: 28.4 ± 4.1; Diabetes n/a; Hypertension: n/a | **Continuous stationery cycling:**  **Condit. Intensity:** Yes (No info.)  **Final intensity:** Moderate-to-Vigorous; 70-80% HR_max_  **Monit. Device:** No info.  **Volume:** Total duration (45 min); Stimuli duration (30 min); 2 to 3 sess/week for 12 weeks ^§^  **Resistance and balance exercise:**  **Volume:** Total duration (No info.); Stimuli duration (No info.); 2 to 3sess/week; 12 weeks ^§^  **No treatment:**  **Volume:** 12 weeks | Pain intensity (NRS average perceived last 2 weeks) | No differences found between aerobic and no treatment group  No differences found between aerobic and resistance group | | | Mod-Vig AE ≈ No Treatment  Mod-Vig AE ≈ Resistance |
| Salacinski et al., 2012  *Parallel RCT* | **KOA diagnostic criteria:** Clinical and radiological according to KL criteria *(grades I-III)*  **Presence of pain:** Yes, pain on the previous month | **Moderate AE** (n_allo_=19, n_an_=19):  Age: 55.1 ± 10.5; F (n=15); BMI: 22.4 ±3.3; Diabetes n/a; Hypertension: n/a  **No treatment** (n_allo_=18, n_an_=18):  Age: 60.6 ± 8.4; F (n=12); BMI: 25.7 ± 6.3; Diabetes n/a; Hypertension: n/a | **Continuous stationary cycling:**  **Condit. Intensity:** No  **Final intensity:** Moderate-to-Vigorous; 70-75% HR_max_  **Monit. Device:** Polar Electro Inc, Lake Success, NY  **Volume:** Total duration (40 progressed to 60 min); Stimuli duration (No info.); 2 sess/week for 12 weeks  **No treatment:**  **Volume:** 12 weeks | Pain intensity (VAS at rest and after 6-min normal walking) | No differences between groups after intervention for pain at rest  Aerobic group showed greater reduction than the no treatment group after 6 min of normal walking | | | Mod-Vig AE ≈ No Treatment  Mod-Vig AE < No Treatment |
|  |  |  |  | Walking performance (normal walk speed in 3.66m, maximal walk speed at 3.66m) | Aerobic group showed greater normal walk speed than the no treatment group  No differences found between groups for maximal walk speed | | | Mod-Vig AE > No Treatment  Mod-Vig AE ≈ No Treatment |
|  |  |  |  | Stiffness (WOMAC- Stiffness) | Aerobic group showed stiffness than the no treatment group | | | Mod-Vig AE < Control |
|  |  |  |  | Disability associated with KOA (WOMAC- Total) | No differences found between aerobic and no treatment group | | | Mod-Vig AE ≈ Control |
| Samut et al., 2015  *Parallel RCT* | **KOA diagnostic criteria:** Clinical and radiological criteria according to ACR *(No specific reference)*  **Presence of pain:**  Probable but unclear | **Vigorous AE** (n_allo_=14, n_an_=14):  Age: 57.57 ± 5.79; F: n/a; BMI: 33.94 ± 7.33; Diabetes n/a; Hypertension: n/a  **Resistance exercise** (n_allo_=15, n_an_=13):  Age: 62.46 ± 7.71; F: n/a; BMI: 30.54 ± 4.45; Diabetes n/a; Hypertension: n/a  **No treatment** (n_allo_=13, n_an_=13):  Age: 60.92 ± 8.85; F: n/a; BMI: 30.36 ± 5.67; Diabetes n/a; Hypertension: n/a | **Vigorous stationary cycling:**  **Condit. Intensity:** Vigorous; 65-70% HRR  **Final intensity:** Vigorous; 70-75% HRR  **Monit. Device:** No info.  **Volume:** Total duration (No info.); Stimuli duration (No info.); 3 sess/week for 6 weeks  **Resistance exercise:**  **Volume:** Total duration (No info.); Stimuli duration (No info.) 3sess/week; 6 weeks  **Minimal education:**  **Volume:** No info. of the duration | Pain intensity (VAS and WOMAC- Pain) | Aerobic group showed greater reduction than the no treatment group in VAS and WOMAC-Pain  No differences found between aerobic and resistance group in VAS and WOMAC-Pain | | | Vig-AE < No treatment  Vig-AE ≈ Resistance |
|  |  |  |  | Walking performance (6MWT) | No differences found between aerobic and no treatment groups  No differences found between aerobic and resistance groups | | | Vig-AE ≈ No Treatment  Vig-AE ≈ Resistance |
|  |  |  |  | Function in sit to stand (30s Chair-to-Stand) | No differences found between aerobic and no treatment groups  No differences found between aerobic and resistance groups | | | Vig-AE ≈ No Treatment  Vig-AE ≈ Resistance |
|  |  |  |  | Stiffness (WOMAC- Stiffness) | No differences found between aerobic and no treatment groups  No differences found between aerobic and resistance groups | | | Vig-AE ≈ No Treatment  Vig-AE ≈ Resistance |
|  |  |  |  | Disability associated with KOA (WOMAC- Total) | Greater reduction in disability in aerobic compared to no treatment group  No differences found between aerobic and resistance groups | | | Vig-AE < No Treatment  Vig-AE ≈ Resistance |
| Watanabe & Someya 2013  *Parallel RCT* | **KOA diagnostic criteria:** Clinical (JOA score) and radiological (YCCS)  **Presence of pain:** Probable, as it was considered as a component of clinical criteria | **Very light-to-light AE** (n_allo_=15, n_an_=12):  Age: 75 ± 7.6; F (n= 10); BMI: 22.7 ±3.1; Diabetes n/a; Hypertension: n/a  **Very light-to-light AE** (n_allo_=15, n_an_=13):  Age: 80 ± 5.9; F (n= 11); BMI: 22.2 ± 3.7; Diabetes n/a; Hypertension: n/a | **Full-body-weight continuous walking:**  **Condit. Intensity:** No  **Final intensity:** Very light-to-light; 40-60% HR_max_ and 12-14 Borg (employed both parameters)  **Monit. Device:** Poral RS100  **Volume:** Total duration (28 min); Stimuli duration (20 min); 2 sess/week for 6 weeks  **Body-weight-supported continuous walking:**  **Condit. Intensity:** No  **Final intensity:** Very light-to-light ;40-60% HR_max_ or 12-14 Borg employed both parameters)  **Monit. Device:** Poral RS100  **Volume:** Total duration (28 min); Stimuli duration (20 min); 2 sess/week for 6 weeks | Pain intensity (VAS) | Greater reduction in body-weight supported AE compared to full body weight group | | | Very light-light AE Full body weight > Very light-light Partial weight |
|  |  |  |  | Walking performance (6MWT (m) and 10 m WT (s)) | Greater distance in 6MWT and greater speed in 10m WT in the body weight supported group compared to full weight group | | | Very light-light Partial weight > Very light-light Full body weight |
|  |  |  |  | Function in sit to stand and walking (TUG) | No differences found between full weight and body weight supported groups | | | Very light-light Full body weight ≈ Very light-light Partial weight |

6MWT, 6-minute walking test; 10mWT, 10-meter walking test; ACR, American College of rheumatology; AE, Aerobic exercise; BMI, Body mass index; BPI, Brief Pain inventory; %HRR, Heart rate reserve; JOA, Japan Orthopedic Association score; KL, Kellgren–Lawrence radiological OA criteria; KOA, Knee osteoarthritis; Mod, Moderate; n_alo_, sample allocated; n_an_, sample analysed; NRS, Numeric Rating Scale; VAS, Visual analogic scale; Vig, Vigorous; WOMAC, Western Ontario and McMaster Universities Osteoarthritis index; YCC, Yokohama City Classification System; %HR_max_, % Theoretical Maximum Heart Rate .

* analysed sample was presented as a whole not-divided-by group. Therefore, for meta-analysis it was assumed to be the 50% of the sample.

^†^ number of weeks was estimated from the value of 18 months. This was determined by computing the total number of days along a 1.5 years (≈547.5) and dividing them by 7 days (78.2 weeks).

^§^ weekly frequency assumed to be 2.5 for meta-analysis.
