## Supplementary material for "How intense is effective? Exploring aerobic exercise intensity for knee osteoarthritis through a Bayesian network meta-analysis": T2 Data availability

Table 2. Data availability, extraction and estimations for meta-analyses.

| **Outcome measure** | **Analyses outcome measure of interest (k)** | **Original studies (k)** | **Comparisons eligible for meta-analysis (k)** | **Text/Table or Plot (k)** | **Included in the meta-analysis** | | |
| --- | --- | --- | --- | --- | --- | --- | --- |
|  |  |  |  |  | **Extractable (k)** | **Raw extraction as Mean and SD (k)** | **Instruments employed in NMA (k)** |
| **Pain intensity** | Yes (12): (Arrieiro et al., 2019; Beckwée et al., 2015; Casilda-López et al., 2017; de Almeida et al., 2019, 2020; Ettinger et al., 1997; Lim et al., 2010; Messier et al., 1997; Øiestad et al., 2023; Salacinski et al., 2012; Samut et al., 2015; Watanabe & Someya, 2013). | Yes (10): (Arrieiro et al., 2019; Beckwée et al., 2015; Casilda-López et al., 2017; de Almeida et al., 2019; Ettinger et al., 1997; Lim et al., 2010; Øiestad et al., 2023; Salacinski et al., 2012; Samut et al., 2015; Watanabe & Someya, 2013). | Yes (7): (Beckwée et al., 2015; de Almeida et al., 2019; Ettinger et al., 1997; Lim et al., 2010; Øiestad et al., 2023; Salacinski et al., 2012; Samut et al., 2015). | Text/Table (7): (Beckwée et al., 2015; de Almeida et al., 2019; Ettinger et al., 1997; Lim et al., 2010; Øiestad et al., 2023; Salacinski et al., 2012; Samut et al., 2015). | Yes (7): (Beckwée et al., 2015; de Almeida et al., 2019; Ettinger et al., 1997; Lim et al., 2010; Øiestad et al., 2023; Salacinski et al., 2012; Samut et al., 2015). | Yes (5): (de Almeida et al., 2019; Lim et al., 2010; Øiestad et al., 2023; Salacinski et al., 2012; Samut et al., 2015).  No (2): Mean estimated from median**^⁑^** (Beckwée et al., 2015); SD estimated from IQR**^⁂^** (Ettinger et al., 1997). | VAS (2): (de Almeida et al., 2019; Samut et al., 2015)  VAS after 6-min normal walking (1): (Salacinski et al., 2012) |
|  |  |  |  |  |  |  | NRS maximal the previous 24h (1): (Beckwée et al., 2015)  NRS average the previous 2 weeks (1): (Øiestad et al., 2023) |
|  |  |  |  |  |  |  | BPI (1): (Lim et al., 2010). |
|  |  |  |  |  |  |  | 1-6 scale (1): (Ettinger et al., 1997). |
|  |  |  | No (3): Exclusive comparisons of groups with equal aerobic exercise intensities (Arrieiro et al., 2019; Casilda-López et al., 2017; Watanabe & Someya, 2013). |  |  |  |  |
|  |  | No (2): Subsample of Ettinger et al., 2019 with fewer subjects (Messier et al., 1997); Same sample as de Almeida et al., 2019 (de Almeida et al., 2020)**^*^** |  |  |  |  |  |
|  | No (3): (Bavardi Moghadam & Shojaedin, 2017; Keogh et al., 2018; Mangione et al., 1999). |  |  |  |  |  |  |
| **Function in walking tasks** | Yes (11): (Arrieiro et al., 2019; Bavardi Moghadam & Shojaedin, 2017; Casilda-López et al., 2017; de Almeida et al., 2020; Ettinger et al., 1997; Keogh et al., 2018; Mangione et al., 1999; Messier et al., 1997; Salacinski et al., 2012; Samut et al., 2015; Watanabe & Someya, 2013). | Yes (10): (Arrieiro et al., 2019; Bavardi Moghadam & Shojaedin, 2017; Casilda-López et al., 2017; de Almeida et al., 2020; Ettinger et al., 1997; Keogh et al., 2018; Mangione et al., 1999; Salacinski et al., 2012; Samut et al., 2015; Watanabe & Someya, 2013). | Yes (7): (Bavardi Moghadam & Shojaedin, 2017; de Almeida et al., 2020; Ettinger et al., 1997; Keogh et al., 2018; Mangione et al., 1999; Salacinski et al., 2012; Samut et al., 2015). | Text/Table (7): (Bavardi Moghadam & Shojaedin, 2017; de Almeida et al., 2020; Ettinger et al., 1997; Keogh et al., 2018; Mangione et al., 1999; Salacinski et al., 2012; Samut et al., 2015). | Yes (7): (Bavardi Moghadam & Shojaedin, 2017; de Almeida et al., 2020; Ettinger et al., 1997; Keogh et al., 2018; Mangione et al., 1999; Salacinski et al., 2012; Samut et al., 2015). | Yes (6): (Bavardi Moghadam & Shojaedin, 2017; de Almeida et al., 2020; Keogh et al., 2018; Mangione et al., 1999; Salacinski et al., 2012; Samut et al., 2015).  No (1): SD estimated from SE^‡^ (Ettinger et al., 1997) | 6MWT (4) (m): (Bavardi Moghadam & Shojaedin, 2017; Ettinger et al., 1997; Mangione et al., 1999; Samut et al., 2015). |
|  |  |  |  |  |  |  | Normal walk speed in 3.66m (2) (m/s): (Keogh et al., 2018; Salacinski et al., 2012). |
|  |  |  |  |  |  |  | 40m walk test (1) (m/s): (de Almeida et al., 2020). |
|  |  |  | No (3): Exclusive comparisons of groups with equal aerobic exercise intensities (Arrieiro et al., 2019; Casilda-López et al., 2017; Watanabe & Someya, 2013). |  |  |  |  |
|  |  | No (1): Subsample of Ettinger et al., 2019 with fewer subjects (Messier et al., 1997) |  |  |  |  |  |
|  | No (4): (Beckwée et al., 2015; de Almeida et al., 2019; Lim et al., 2010; Øiestad et al., 2023). |  |  |  |  |  |  |

NMA, network meta-analysis; VAS, visual analogue scale; NPRS, numerical pain rating scale; BPI, brief pain intensity scale; 6MWT, 6 minute walk test;

**^*^** Although both studies by de Almeida et al. (2019, 2020) analyzed pain intensity using the same sample, they employed different instruments (VAS in 2019; WOMAC-Pain in 2020). Data from the 2019 study was selected due to the higher validity and precision of the VAS for estimating pain intensity.

**^⁑^**	Median to Mean was calculated using the formula number 14 by Wan et al., 2014$: \left[ Mean\approx\frac{Q1+Median+Q3}{3} \right]$. Q3 and Q1 were estimated as having the same distance from the median.

**^⁂^** IQR to SD was calculated using the following formula number 15 by Wan et al., 2014: $\left[ SD\approx\frac{Q3-Q1}{\eta(n)} \right]$

^‡^ SE to SD was calculated using the following formula: $\left[ SD\approx\sqrt{n} \times SE \right]$

6MWT, 6-minute walking test; BPI, brief pain inventory; NRS, numerical rating scale; VAS, visual analogue scale; WOMAC-Pain, Western Ontario and McMaster Universities Osteoarthritis index pain subscale;
