## Supplementary material for "How intense is effective? Exploring aerobic exercise intensity for knee osteoarthritis through a Bayesian network meta-analysis": T3 GREADE Pain

Table 3. Summary Grading of Evidence for network meta-analysis for different intensities of aerobic exercise in KOA on pain intensity.

| **Outcome** | **Comparison** | | **Step Analysis** | **Evidence** | | **Risk of bias** | **Indirectness** | **Imprecision** | | | **Heterogeneity** | | **Pub. bias** | **Intransitivity** | **Incoherence** | | **Cert** |
| --- | --- | --- | --- | --- | --- | --- | --- | --- | --- | --- | --- | --- | --- | --- | --- | --- | --- |
|  | **Group 1** | **Group 2** |  |  |  |  |  | **Model** | **Hedges’ *g* (95%CrI)** | **Rating** | **Result** | **Rating** |  |  | **Z p-value** | **Rating** |  |
| Pain intensity | Light-Moderate-Vigorous | Moderate | Preliminary analysis | Direct |  | n.a | n.a | n.a | n.a | - | n.a | n.a | n.a | - | - | - | n.a |
|  |  |  |  | Indirect | 2^nd^ order loop: LMV vs RE vs NT vs M | Very serious (‒2) | Not serious (0) | Model 1 | ‒0.38 (‒4.88, 4.12) | - | τ = 1.4 | Very serious (‒2) | Very serious (‒2) | Not serious (0) | - | - | Very low |
|  |  |  |  |  |  |  |  | Model 2 | ‒0.32 (‒7.49, 6.84) | - | τ = 1.4 | Very serious (‒2) | Very serious (‒2) | Not serious (0) | - | - |  |
|  |  |  | Final analysis | Mixed | - | - | - | Model 1 | ‒0.38 (‒4.88, 4.12) | Very serious (‒2) | - | - | - | - | n.a | n.a | Very low |
|  |  |  |  |  |  |  |  | Model 2 | ‒0.32 (‒7.49, 6.84) | Very serious (‒2) |  |  |  |  | n.a | n.a |  |
|  | Light-Moderate-Vigorous | Moderate-Vigorous | Preliminary analysis | Direct |  | n.a | n.a | n.a | n.a | - | n.a | n.a | n.a | n.a | - | - | n.a |
|  |  |  |  | Indirect | 1^st^ order loop: LMV vs RE vs MV | Very serious (‒2) | Not serious (0) | Model 1 | ‒0.15 (‒3.65, 3.34) | - | τ = 1.4 | Very serious (‒2) | Serious (‒1) | Not serious (0) | - | - | Very low |
|  |  |  |  |  |  |  |  | Model 2 | ‒0.15 (‒3.69, 3.38) | - | τ = 1.4 | Very serious (‒2) | Serious (‒1) | Not serious (0) | - | - |  |
|  |  |  | Final analysis | Mixed | - | - | - | Model 1 | ‒0.15 (‒3.65, 3.34) | Very serious (‒2) | - | - | - | - | n.a | n.a | Very low |
|  |  |  |  |  |  |  |  | Model 2 | ‒0.15 (‒3.69, 3.38) | Very serious (‒2) |  |  |  |  | n.a | n.a |  |
|  | Light-Moderate-Vigorous | Vigorous | Preliminary analysis | Direct |  | n.a | n.a | n.a | n.a | - | n.a | n.a | n.a | n.a | - | - | n.a |
|  |  |  |  | Indirect | 1^st^ order loop: LMV vs RE vs V | Very serious (‒2) | Not serious (0) | Model 1 | 0.27 (‒3.32, 3.85) | - | τ = 1.4 | Very serious (‒2) | Serious (‒1) | Not serious (0) | - | - | Very low |
|  |  |  |  |  |  |  |  | Model 2 | 0.27 (‒3.35, 3.89) | - | τ = 1.4 | Very serious (‒2) | Serious (‒1) | Not serious (0) | - | - |  |
|  |  |  | Final analysis | Mixed | - | - | - | Model 1 | 0.27 (‒3.32, 3.85) | Very serious (‒2) | - | - | - | - | n.a | n.a | Very low |
|  |  |  |  |  |  |  |  | Model 2 | 0.27 (‒3.35, 3.89) | Very serious (‒2) |  |  |  |  | n.a | n.a |  |
|  | Moderate | Moderate-Vigorous | Preliminary analysis | Direct |  | n.a | n.a | n.a | n.a | - | n.a | n.a | n.a | n.a | - | - | n.a |
|  |  |  |  | Indirect | 1^st^ order loop: M vs NT vs MV | Very serious (‒2) | Not serious (0) | Model 1 | 0.23 (‒3.76, 4.21) | - | τ = 1.4 | Very serious (‒2) | Not serious (0) | Not serious (0) | - | - | Very low |
|  |  |  |  |  |  |  |  | Model 2 | 0.17 (‒6.67, 7.00) | - | τ = 1.4 | Very serious (‒2) | Not serious (0) | Not serious (0) | - | - |  |
|  |  |  | Final analysis | Mixed | - | - | - | Model 1 | 0.23 (‒3.76, 4.21) | Very serious (‒2) | - | - | - | - | n.a | n.a | Very low |
|  |  |  |  |  |  |  |  | Model 2 | 0.17 (‒6.67, 7.00) | Very serious (‒2) |  |  |  |  | n.a | n.a |  |
|  | Moderate | Vigorous | Preliminary analysis | Direct |  | n.a | n.a | n.a | n.a | - | n.a | n.a | n.a | n.a | - | - | n.a |
|  |  |  |  | Indirect | 1^st^ order loop: M vs NT vs V | Very serious (‒2) | Not serious (0) | Model 1 | 0.65 (‒3.42, 4.71) | - | τ = 1.4 | Very serious (‒2) | Serious (‒1) | Not serious (0) | - | - | Very low |
|  |  |  |  |  |  |  |  | Model 2 | 0.59 (‒6.29, 7.47) | - | τ = 1.4 | Very serious (‒2) | Serious (‒1) | Not serious (0) | - | - |  |
|  |  |  | Final analysis | Mixed | - | - | - | Model 1 | 0.65 (‒3.42, 4.71) | Very serious (‒2) | - | - | - | - | n.a | n.a | Very low |
|  |  |  |  |  |  |  |  | Model 2 | 0.59 (‒6.29, 7.47) | Very serious (‒2) |  |  |  |  | n.a | n.a |  |
|  | Moderate-Vigorous | Vigorous | Preliminary analysis | Direct |  | n.a | n.a | n.a | n.a | - | n.a | n.a | n.a | n.a | - | - | n.a |
|  |  |  |  | Indirect | 1^st^ order loop: MV vs RE vs V | Very serious (‒2) | Not serious (0) | Model 1 | 0.42 (‒2.49, 3.33) | - | τ = 1.4 | Very serious (‒2) | Serious (‒1) | Not serious (0) | - | - | Very low |
|  |  |  |  |  |  |  |  | Model 2 | 0.43 (‒2.49, 3.34) | - | τ = 1.4 | Very serious (‒2) | Serious (‒1) | Not serious (0) | - | - |  |
|  |  |  | Final analysis | Mixed | - | - | - | Model 1 | 0.42 (‒2.49, 3.33) | Very serious (‒2) | - | - | - | - | n.a | n.a | Very low |
|  |  |  |  |  |  |  |  | Model 2 | 0.43 (‒2.49, 3.34) | Very serious (‒2) |  |  |  |  | n.a | n.a |  |

Cert, certainty; LMV, light-to-moderate-to-vigorous aerobic exercise; M, moderate aerobic exercise; MV, moderate-to-vigorous aerobic exercise; n.a., not analysed; NT, no treatment; RE, resistance exercise; V, virogorous aerobic exercise: Pub. Bias, publication bias;
