## Supplementary material for "How intense is effective? Exploring aerobic exercise intensity for knee osteoarthritis through a Bayesian network meta-analysis": T4 GRADE Walking

Table 4. Summary Grading of Evidence for network meta-analysis for different intensities of aerobic exercise in KOA on walking performance.

| **Outcome** | **Comparison** | | **Step Analysis** | **Evidence** | | **Risk of bias** | **Indirectness** | **Imprecision** | | | **Heterogeneity** | | **Pub. bias** | **Intransitivity** | **Incoherence** | | **Cert** |
| --- | --- | --- | --- | --- | --- | --- | --- | --- | --- | --- | --- | --- | --- | --- | --- | --- | --- |
|  | **Group 1** | **Group 2** |  |  |  |  |  | **Model** | **Hedges’ *g* (95%CrI)** | **Rating** | **Result** | **Rating** |  |  | **Z p-value** | **Rating** |  |
| Walking performance | Light-Moderate | Light-Moderate-Vigorous | Preliminary analysis | Direct |  | n.a | n.a | n.a | n.a | - | n.a | n.a | n.a | - | - | - | n.a |
|  |  |  |  | Indirect | 2^nd^ order loop: LM vs V vs RE vs LMV | Very serious (‒2) | Not serious (0) | Model 1 | 0.44 (‒3.42, 4.30) | - | τ = 0.89 | Very serious (‒2) | Serious (‒1) | Not serious (0) | - | - | Low |
|  |  |  |  |  |  |  |  | Model 2 | ‒0.08 (‒2.23, 2.07) | - | τ = 0.30 | Not serious (0) | Not serious (0) | Not serious (0) | - | - |  |
|  |  |  | Final analysis | Mixed | - | - | - | Model 1 | 0.44 (‒3.42, 4.30) | Very serious (‒2) | - | - | - | - | n.a | n.a | Very low |
|  |  |  |  |  |  |  |  | Model 2 | ‒0.08 (‒2.23, 2.07) | Very serious (‒2) |  |  |  |  | n.a | n.a |  |
|  | Light-Moderate | Moderate | Preliminary analysis | Direct |  | n.a | n.a | n.a | n.a | - | n.a | n.a | n.a | n.a | - | - | n.a |
|  |  |  |  | Indirect | 1^st^ order loop: LM vs V vs M | Very serious (‒2) | Not serious (0) | Model 1 | 0.11 (‒3.52, 3.74) | - | τ = 0.89 | Very serious (‒2) | Serious (‒1) | Not serious (0) | - | - | Low |
|  |  |  |  |  |  |  |  | Model 2 | ‒0.27 (‒2.74, 2.21) | - | τ = 0.30 | Not serious (0) | Not serious (0) | Not serious (0) | - | - |  |
|  |  |  | Final analysis | Mixed | - | - | - | Model 1 | 0.11 (‒3.52, 3.74) | Very serious (‒2) | - | - | - | - | n.a | n.a | Very low |
|  |  |  |  |  |  |  |  | Model 2 | ‒0.27 (‒2.74, 2.21) | Very serious (‒2) |  |  |  |  | n.a | n.a |  |
|  | Light-Moderate | Moderate-Vigorous | Prelimiry analysis | Direct |  | n.a | n.a | n.a | n.a | - | n.a | n.a | n.a | n.a | n.a |  | n.a |
|  |  |  |  | Indirect | 2^nd^ order loop: LM vs V vs RE vs MV | Very serious (‒2) | Not serious (0) | Model 1 | 1.04 (‒2.68, 4.76) | - | τ = 0.89 | Very serious (‒2) | Serious (‒1) | Not serious (0) |  |  | Low |
|  |  |  |  |  |  |  |  | Model 2 | 0.12 (‒1.97, 2.22) | - | τ = 0.30 | Not serious (0) | Not serious (0) | Not serious (0) |  |  |  |
|  |  |  | Final analysis | Mixed | - | - | - | Model 1 | 1.04 (‒2.68, 4.76) | Very serious (‒2) | - | - | - | - | n.a | n.a | Very low |
|  |  |  |  |  |  |  |  | Model 2 | 0.12 (‒1.97, 2.22) | Very serious (‒2) |  |  |  |  | n.a | n.a |  |
|  | Light-Moderate | Vigorous | Preliminary analysis | Direct | Mangione 1997 | Very serious (‒2) | Not serious (0) | Model 1 | ‒0.13 (‒0.82, 0.57) | - | n.a | Not serious (0) | Not serious (0) | - | - | - | Low |
|  |  |  |  |  |  |  |  | Model 2 | ‒0.13 (‒0.82, 0.57) | - | n.a | Not serious (0) | Not serious (0) |  |  |  |  |
|  |  |  |  | Indirect | n.a | n.a | n.a | n.a | n.a | - | n.a | n.a | n.a | n.a | n.a | n.a | n.a |
|  |  |  | Final analysis | Mixed | - | - | - | Model 1 | ‒0.13 (‒3.25, 3.51) | Very serious (‒2) | - | - | - | - | n.a | n.a | Very low |
|  |  |  |  |  |  |  |  | Model 2 | 0.13 (‒1.74, 1.99) | Very serious (‒2) |  |  |  |  | n.a | n.a |  |
|  | Light-Moderate-Vigorous | Moderate | Preliminary analysis | Direct |  | n.a | n.a | n.a | n.a | - | n.a | n.a | n.a | n.a | n.a |  | n.a |
|  |  |  |  | Indirect | 2^nd^ order loop: LMV vs RE vs NT vs M | Very serious (‒2) | Not serious (0) | Model 1 | ‒0.33 (‒3.51, 2.86) | - | τ = 0.89 | Very serious (‒2) | Very serious (‒2) | Not serious (0) | - | - | Very low |
|  |  |  |  |  |  |  |  | Model 2 | ‒0.19 (‒2.50, 2.13) | - | τ = 0.30 | Not serious (0) | Very serious (‒2) | Not serious (0) | - | - |  |
|  |  |  | Final analysis | Mixed | - | - | - | Model 1 | ‒0.33 (‒3.51, 2.86) | Very serious (‒2) | - | - | - | - | n.a | n.a | Very low |
|  |  |  |  |  |  |  |  | Model 2 | ‒0.19 (‒2.50, 2.13) | Very serious (‒2) |  |  |  |  | n.a | n.a |  |
|  | Light-Moderate-Vigorous | Moderate-Vigorous | Preliminary analysis | Direct |  | n.a | n.a | n.a | n.a | - | n.a | n.a | n.a | n.a | n.a |  | n.a |
|  |  |  |  | Indirect | 1^st^ order loop: LMV vs RE vs MV | Very serious (‒2) | Not serious (0) | Model 1 | 0.60 (‒2.69, 3.90) | - | τ = 0.89 | Very serious (‒2) | Not serious (0) | Not serious (0) | - | - | Low |
|  |  |  |  |  |  |  |  | Model 2 | 0.20 (‒1.70, 2.10) | - | τ = 0.30 | Not serious (0) | Not serious (0) | Not serious (0) |  |  |  |
|  |  |  | Final analysis | Mixed | - | - | - | Model 1 | 0.60 (‒2.69, 3.90) | Very serious (‒2) | - | - | - | - | n.a | n.a | Very low |
|  |  |  |  |  |  |  |  | Model 2 | 0.20 (‒1.70, 2.10) | Very serious (‒2) |  |  |  |  | n.a | n.a |  |
|  | Light-Moderate-Vigorous | Vigorous | Preliminary analysis | Direct |  | n.a | n.a | n.a | n.a | - | n.a | n.a | n.a | n.a | n.a |  | n.a |
|  |  |  |  | Indirect | 1^st^ order loop: LMV vs RE vs V | Very serious (‒2) | Not serious (0) | Model 1 | ‒0.31 (‒3.21, 2.60) | - | τ = 0.89 | Very serious (‒2) | Serious (‒1) | Not serious (0) | - | - | Low |
|  |  |  |  |  |  |  |  | Model 2 | 0.21 (‒1.44, 1.85) | - | τ = 0.30 | Not serious (0) | Not serious (0) | Not serious (0) |  |  |  |
|  |  |  | Final analysis | Mixed | - | - | - | Model 1 | ‒0.31 (‒3.21, 2.60) | Very serious (‒2) | - | - | - | - | n.a | n.a | Very low |
|  |  |  |  |  |  |  |  | Model 2 | 0.21 (‒1.44, 1.85) | Very serious (‒2) |  |  |  |  | n.a | n.a |  |
|  | Moderate | Moderate-Vigorous | Preliminary analysis | Direct |  | n.a | n.a | n.a | n.a | - | n.a | n.a | n.a | n.a | n.a |  | n.a |
|  |  |  |  | Indirect | 1^st^ order loop: M vs NT vs MV | Very serious (‒2) | Not serious (0) | Model 1 | 0.93 (‒2.10, 3.95) | - | τ = 0.89 | Very serious (‒2) | Very serious (‒2) | Not serious (0) | - | - | Very low |
|  |  |  |  |  |  |  |  | Model 2 | 0.39 (‒1.87, 2.65) | - | τ = 0.30 | Not serious (0) | Very serious (‒2) | Not serious (0) |  |  |  |
|  |  |  | Final analysis | Mixed | - | - | - | Model 1 | 0.93 (‒2.10, 3.95) | Very serious (‒2) | - | - | - | - | n.a | n.a | Very low |
|  |  |  |  |  |  |  |  | Model 2 | 0.39 (‒1.87, 2.65) | Very serious (‒2) |  |  |  |  | n.a | n.a |  |
|  | Moderate | Vigorous | Preliminary analysis | Direct | Keogh 2018 | Very serious (‒2) | Not serious (0) | Model 1 | ‒0.39 (‒0.57, 1.35) | - | n.a | Not serious (0) | Not serious (0) | - | - | - | Low |
|  |  |  |  |  |  |  |  | Model 2 | ‒0.39 (‒0.57, 1.35) | - | n.a | Not serious (0) | Not serious (0) | - | - | - | Low |
|  |  |  |  | Indirect | 1^st^ order loop: M vs NT vs V | Very serious (‒2) | Not serious (0) | Model 1 | ‒0.26 (‒3.97, 3.47) | - | τ = 1.41 | Very serious (‒2) | Serious (‒1) | Not serious (0) | - | - | Very low |
|  |  |  |  |  |  |  |  | Model 2 | ‒0.28 (‒3.97, 3.42) | - | τ = 1.40 | Very serious (‒2) | Very serious (‒2) | Not serious (0) |  |  |  |
|  |  |  | Final analysis | Mixed | - | - | - | Model 1 | 0.02 (‒2.57, 2.61) | Very serious (‒2) | - | - | - | - | 0.74 | Not serious (0) | Very low |
|  |  |  |  |  |  |  |  | Model 2 | 0.40 (‒1.66, 2.44) | Very serious (‒2) |  |  |  |  | 0.73 | Not serious (0) |  |
|  | Moderate-Vigorous | Vigorous | Preliminary analysis | Direct |  | n.a | n.a | n.a | n.a | - | n.a | n.a | n.a | n.a | n.a |  | n.a |
|  |  |  |  | Indirect | 1^st^ order loop: MV vs NT vs V | Very serious (‒2) | Not serious (0) | Model 1 | ‒0.91 (‒3.63, 1.81) | - | τ = 0.89 | Very serious (‒2) | Serious (‒1) | Not serious (0) | - | - | Low |
|  |  |  |  |  |  |  |  | Model 2 | 0.00 (‒1.56, 1.57) | - | τ = 0.30 | Not serious (0) | Not serious (0) | Not serious (0) |  |  |  |
|  |  |  | Final analysis | Mixed | - | - | - | Model 1 | ‒0.91 (‒3.63, 1.81) | Very serious (‒2) | - | - | - | - | n.a | n.a | Very low |
|  |  |  |  |  |  |  |  | Model 2 | 0.00 (‒1.56, 1.57) | Very serious (‒2) |  |  |  |  |  |  |  |

Cert, certainty; LM, light-to-moderate; LMV, light-to-moderate-to-vigorous aerobic exercise; M, moderate aerobic exercise; MV, moderate-to-vigorous aerobic exercise; n.a., not analysed; NT, no treatment; RE, resistance exercise; V, virogorous aerobic exercise: Pub. Bias, publication bias;
